# In-depth mass-spectrometry reveals phospho-RAB12 as a blood biomarker of G2019S LRRK2-driven Parkinson’s

**DOI:** 10.1101/2024.05.04.24306824

**Authors:** Adriana Cortés, Toan K. Phung, Lorena de Mena, Alicia Garrido, Jon Infante, Javier Ruíz-Martínez, Miquel À. Galmés-Ordinas, Sophie Glendinning, Jesica Pérez, Ana Roig, Marta Soto, Marina Cosgaya, Valeria Ravasi, Manel Fernández, Alejandro Rubiano-Castro, Ramón Díaz, Haizea Hernández-Eguiazu, Coro Sánchez-Quintana, Ana Vinagre-Aragón, Elisabet Mondragón, Ioana Croitoru, María Rivera-Sánchez, Andrea Corrales-Pardo, María Sierra, Eduardo Tolosa, Cristina Malagelada, Raja S. Nirujogi, Joaquín Fernández-Irigoyen, Enrique Santamaría, Dario R. Alessi, María J. Martí, Mario Ezquerra, Rubén Fernández-Santiago

## Abstract

Leucine-rich repeat kinase 2 (LRRK2) inhibition is a promising disease-modifying therapy for LRRK2-associated Parkinson’s disease (L2PD) and idiopathic PD (iPD). Yet, pharmaco-dynamic readouts and progression biomarkers for disease modification clinical trials are insufficient. Employing phospho-/proteomic analyses we assessed the impact that LRRK2 activating mutations had in peripheral blood mononuclear cells (PBMCs) from a LRRK2 clinical cohort from Spain (n=174) encompassing G2019S L2PD patients (n=37), non-manifesting LRRK2 mutation carriers of G2019S, here, G2019S L2NMCs (n=27), R1441G L2PD patients (n=14), R1441G L2NMCs (n=11), iPD (n=40), and controls (n=45). We identified 207 differential proteins in G2019S L2PD compared to controls (39 up/ 168 down) and 67 in G2019S L2NMCs (10 up/ 57 down). G2019S down-regulated proteins affected the endolysosomal pathway, proteostasis and mitochondria, e.g., ATIC, RAB9A, or LAMP1. At the phospho-proteome level, we observed increases in endogenous phosphorylation levels of pSer106 RAB12 in G2019S carriers, which were validated by immunoblotting after 1 year of follow-up (n=48). Freshly collected PBMCs from 3 G2019S L2PD, 1 R1441G L2PD, 1 iPD, and 5 controls (n=10) showed strong diminishment of pSer106 RAB12 phosphorylation levels after in-vitro administration of the MLi-2 LRRK2 inhibitor. Using machine learning, we identified an 18-feature G2019S phospho-/protein signature capable of discriminating G2019S L2PD, L2NMCs, and controls with 96% accuracy that correlated with disease severity, i.e., UPDRS-III motor scoring. Our study identified pSer106 RAB12 as an endogenous biomarker in easily accessible PBMCs from G2019S carriers and suggests that phospho-/proteomic findings in human PBMCs such as pSer106 RAB12 can be deployed as a universal pharmaco-dynamic readout for L2PD, L2NMCs, and iPD. Future work may determine whether pSer106 RAB12 could help with patient enrichment and monitoring drug efficacy in LRRK2 clinical trials.

## INTRODUCTION

Activating mutations in the leucine-rich repeat kinase 2 (*LRRK2*), e.g., G2019S or R1441G, increase LRRK2 kinase activity^1–4^ causing autosomal-dominant LRRK2 Parkinson’s (L2PD).^5,6^ By converging pathways, LRRK2 kinase activity appears to be also enhanced in patients with idiopathic PD (iPD),^7–9^ undistinguishable from L2PD at the clinical level.^10,11^ Thus, ongoing clinical trials of small-molecule type-I inhibitors targeting active LRRK2 protein conformation is a promising disease-modifying strategy for a broad spectrum of patients.^12,13^ Moreover, non-manifesting LRRK2 mutation carriers (L2NMCs) are at high risk of PD in an age-dependent progressive manner,^14–16^ representing a candidate population for the continued clinical follow-up and disease course modification by early neuroprotective interventions when needed.^13^

A subset of G-proteins from the Ras-related small GTPase superfamily^17^ was reported as phosphorylation substrates of the LRRK2 Ser/Thr kinase.^2,3^ Among these, pThr73 RAB10 was validated as a LRRK2 substrate^18^ showing elevated phosphorylation levels in a large set of R1441G carriers, symptomatic and asymptomatic, yet not in G2019S,^19^ and also a readout for LRRK2 pharmacological inhibition using Mli-2 or DNL201.^20,21^ Moreover, RAB29^22,23^ and more recently RAB12^24,25^ and RAB32^26^ have been described as key upstream LRRK2 activators. Despite significant progress, we still lack robust pharmaco-dynamic readouts and clinical progression biomarkers useful in disease modification clinical trials.

Employing data-independent acquisition (DIA) mass-spectrometry (MS), we have screened the phospho-/proteome of PBMCs from a LRRK2 clinical cohort (n=174) of G2019S L2PD (n=37), G2019S L2NMCs (n=27), R1441G L2PD (n=14), R1441G L2NMCs (n=11), iPD (n=40), and controls (n=45). We identified differential phospho-/proteins in G2019S and R1441G carriers, symptomatic and asymptomatic. We found elevated levels of pSer106 RAB12 phosphorylation in G2019S carriers. Our results suggest that pSer106 RAB12 comprises an endogenous biomarker in G2019S PBMCs, as it is similarly elevated in both L2PD and L2NMCs. Consistent with RAB12 being phosphorylated by LRRK2 we found that pSer106 RAB12 levels strongly diminished after Mli-2 LRRK2 inhibition in all subjects, regardless of their disease or mutation status. We propose that pSer106 RAB12 could be exploited as a target engagement biomarker in LRRK2 clinical trials.^13^ We provide full open access to all data generated here through FAIR,^27^ and Curtain^28^.

## METHODS

### Subjects

Probands participated in the study after local ethics approval and signed informed consent. Study subjects included LRRK2 mutation carriers, symptomatic and asymptomatic, iPD patients, and healthy controls including healthy spouses and companions of Spanish descent. Patient inclusion criteria were a clinical diagnosis of PD by a movement disorders specialist based on the MDS clinical diagnostic criteria for Parkinson’s.^29^ Exclusion criteria were chronic inflammatory and autoimmune diseases, e.g., Crohn’s (CD), inflammatory bowel disease (IBD), rheumatoid arthritis, systemic lupus erythematosus (SLE); chronic neurological diseases such as myasthenia gravis, chronic use of nonsteroidal anti-inflammatory drugs (NSAIDs) or corticosteroid anti-inflammatory medication, and viral or bacterial infection during the week precedent to blood sample donation. Subjects were recruited at three centres from Spain, Hospital Clínic de Barcelona (n=76) (‘B’),^30^ Hospital Marqués de Valdecilla in Santander (n=55) (‘S’),^31^ and Hospital de Donostia in San Sebastian (n=43) (‘D’)^32^ (**Table 1**). By cohort and subject type, the sample included G2019S L2PD (n=37) (16 from B, 20 from S, and 1 from D), G2019S L2NMCs (n=27) (11 B, 15 S, and 1 D), R1441G L2PD (n=14) (1 B, and 13 D), R1441G L2NMCs (n=11) (3 B, and 8 D), iPD (n=40) (20 B, 10 S, and 10 D), and controls (n=45) (25 B, 10 S, and 10 D). We also collected gender, age at sampling, age-at-onset (AAO), LRRK2 mutation status, kinship to index cases, UPDRS-III,^33^ MoCA,^34^ autoimmune and environmental structured questionnaires, COVID-19 history. Specifically, PD patients had a mean age-at-sampling of 63.5 years for G2019S L2PD, 67.1 for R1441G L2PD, and 67.3 for iPD. Asymptomatic blood relatives of L2PD patients, i.e., L2NMCs, were younger than PD patients with a mean age of 56.7 years for G2019S L2NMCs and 61.1 for R1441G L2NMCs. The AAO was similar for G2019S and R1441G L2PD with 55.1 and 55.8 years respectively, whereas iPD had 62.1 years on average. Mean disease duration was 8.4 years for G2019S L2PD, 12.3 for R1441G L2PD and 5.2 for iPD. Average disease severity, UPDRS-III motor scoring, was similar (mild) in all three patient groups, i.e., 16.0 in G2019S L2PD, 19.8 R1441G L2PD, and 19.7 for iPD. Mean MoCA scores were also mild and similar in all patients, 24.3 for G2019S L2PD, 23.2 for R1441G L2PD, and 25.6 for iPD. As for medication, L-DOPA equivalent daily dose (LEDD) was 635.8 mg for G2019S L2PD, 711.5 mg for R1441G L2PD, and 584.7 mg for iPD.

**Table 1.** Participant clinic-demographics. Data are expressed as a mean ± standard deviation (S.D.) with the number of available subjects/totals in brackets. L2PD = LRRK2-associated PD patients; L2NMC = LRRK2 non-manifesting carriers; iPD = idiopathic PD; C = controls; AAO = age-at-onset; UPDRS-III = Unified Parkinson’s Disease Rating Scale; MoCA = Montreal cognitive assessment; LEDD = levodopa equivalent daily dose; “-“ = not applicable; NA = not available.

| Cohort | N<br>(males/<br>females) | Age at<br>sampling<br>(years) | PD<br>AAO<br>(years) | Disease<br>duration<br>(years) | UPDRS-III | H&Y | MoCA | LEDD<br>(mg) | Passed<br>COVID-19<br>(yes/no) |
| --- | --- | --- | --- | --- | --- | --- | --- | --- | --- |
| **Meta** | **174** |  |  |  |  |  |  |  |  |
| G2019S L2PD | 37 (20/17) | 63.5±9.1 (37/37) | 55.1±10.2 (33/37) | 8.4±6.3 (33/37) | 16.0±9.7 (34/37) | 2.0±0.6 (21/37) | 24.3±4.5 (35/37) | 635.8± 438.8 (29/39) | 4/30 (34/37) |
| G2019S L2NMC | 27 (18/9) | 56.7±14.1 (26/27) | - | - | 1.0±1.6 (22/27) | - | 25.4±6.6 (25/27) | - | 6/19 (25/27) |
| R1441G L2PD | 14 (7/7) | 67.1±9.5 (14/14) | 55.8±11.4 (14/14) | 12.3± 5.5 (14/14) | 19.8±12.0 (14/14) | 2.2± 0.9 (13/14) | 23.2±5.5 (10/14) | 711.5 ± 355.7 (14/14) | 4/10 (14/14) |
| R1441G L2NMC | 11 (4/7) | 61.1±5.5 (11/11) | - | - | 1.2±2.1 (11/11) | - | 28.6±2.0 (11/11) | - | 1/10 (11/11) |
| iPD | 40 (30/10) | 67.3±7.7 (40/40) | 62.1±8.5 (40/40) | 5.2±4.3 (40/40) | 19.7±13.2 (40/40) | 2.2±0.6 (31/40) | 25.6±3.7 (33/40) | 584.7 ± 373.6 (37/40) | 1/37 (38/40) |
| C | 45 (18/27) | 60.0±10.9 (45/45) | - | - | 1.2±2.2 (17/27) | - | 27.5±3.2 (27/27) | - | 14/30 (44/45) |
| **B - Barcelona** | **76** |  |  |  |  |  |  |  |  |
| G2019S L2PD | 16 (7/9) | 65.5± 8.3 (16/16) | 53.5±11.3 (14/16) | 11.4±7.1 (14/16) | 13.0± 7.2 (13/16) | 2.0± 0.5 (11/16) | 25.0±4.3 (14/16) | 596.5 ± 269.7 (13/18) | 2/12 (14/16) |
| G2019S L2NMC | 11 (7/4) | 47.8±15.5 (10/11) | - | - | 0.3±0.7 (10/11) | - | 28.2±2.0 (10/11) | - | 4/5 (9/11) |
| R1441G L2PD | 1 (0/1) | 44.0 (1/1) | 32.0 (1/1) | 12.0 (1/1) | 16.0 (1/1) | NA (0/1) | 30.0 (1/1) | 400 (1/1) | 0/1 (1/1) |
| R1441G L2NMC | 3 (2/1) | 65.3±1.5 (3/3) | - | - | 3.0±3.6 (3/3) | - | 29.3±0.6 (3/3) | - | 1/2 (3/3) |
| iPD | 20 (16/4) | 68.3±7.9 (20/20) | 64.3±7.8 (20/20) | 4.0 ± 3.0 (20/20) | 14.7±4.2 (20/20) | 1.9± 0.2 (20/20) | 27.2± 3.2 (20/20) | 453.8 ± 279.8 (19/20) | 1/19 (20/20) |
| C | 25 (9/16) | 63.9±10.6 (25/25) | - | - | 1.7±2.4 (16/25) | - | 27.9±2.1 (25/25) | - | 8/17 (25/25) |
| **S - Santander** | **55** |  |  |  |  |  |  |  |  |
| G2019S L2PD | 20 (13/0) | 61.3±9.0 (20/20) | 55.6±9.1 (18/20) | 6.1±4.7 (18/20) | 18.1±11.0 (20/20) | 1.9± 0.6 (9/20) | 24.2± 4.6 (20/20) | 652.7 ± 562.3 (15/20) | 2/17 (19/20) |
| G2019S L2NMC | 15 (11/4) | 62.6±10.2 (15/15) | - | - | 1.8±1.9 (11/15) | - | 23.3±8.1 (14/15) | - | 2/13 (15/15) |
| iPD | 10 (6/4) | 67.2±7.6 (10/10) | 62.0±6.3 (10/10) | 5.2±4.0 (10/10) | 17.9±8.3 (10/10) | 2.5 (1/10) | 23±3.5 (10/10) | 479.9 ± 220.2 (8/10) | 0/8 (8/10) |
| C | 10 (4/6) | 56.9±11.1 (10/10) | - | - | NA | - | 25.0±3.6 (6/6) | - | 1/8 (9/10) |
| **D - Donostia** | **43** |  |  |  |  |  |  |  |  |
| G2019S L2PD | 1 (0/1) | 78.0 (1/1) | 68.0 (1/1) | 10.0 (1/1) | 15.0 (1/1) | 3.0 (1/1) | 19.0 (1/1) | 893 (1/1) | 0/1 (1/1) |
| G2019S L2NMC | 1 (0/1) | 58.0 (1/1) | - | - | NA | - | 29 (1/1) | - | 0/1 (1/1) |
| R1441G L2PD | 13 (7/6) | 68.9±7.1 (13/13) | 56.5±9.7 (13/13) | 12.4±5.7 (13/13) | 20.1±12.5 (13/13) | 2.2±0.9 (13/13) | 22.4±5.2 (9/13) | 735.5 ± 358.3 (13/13) | 4/9 (13/13) |
| R1441G L2NMC | 8 (2/6) | 59.5±5.6 (8/8) | - | - | 0.5±0.8 (6/6) | - | 28.3±2.3 (6/6) | - | 0/8 (8/8) |
| iPD | 10 (8/2) | 65.4±7.8 (10/10) | 57.9±10.6 (10/10) | 7.5±5.9 (10/10) | 31.7±20.7 (10/10) | 2.7± 0.7 (10/10) | 23.7±2.9 (10/10) | 917.3 ± 441.8 (10/10) | 0/10 (10/10) |
| C | 10 (5/5) | 53.5±8.1 (10/10) | - | - | NA | - | 29.1±3.9 (5/5) | - | 5/5 (10/10) |

### Genotyping

We genotyped the most common LRRK2 mutations in our population using Taqman SNP assays-on-demand for *LRRK2* G2019S (Thermo Fisher Sci., #C-63498123-10) and a commercial TaqMan assay for *LRRK2* R1441G^35^ on a Step One Plus Real-time PCR System (Life Tech. Inc.)

### PBMC isolation

40 ml of peripheral blood were drawn early in the morning in fasting and PBMCs were isolated by density gradient using Sodium-Citrate tubes (BD Vacutainer CPT, #EAN30382903627821) following manufacturer’s instructions. All samples used in the study were dry PBMC pellets flash-frozen in liquid N_2_ and stored at −80°C for a time period less than a year until use.

### PBMC preparation

PBMC samples from the three cohorts were processed in parallel. Blind experimental groups to the operator were balanced and randomized in runs to avoid manipulation bias. Briefly, PBMCs were homogenized in lysis buffer (7 M urea, 2 M thiourea, 50 mM dithiothreitol) and supplemented with cOmplete Mini protease (Roche, #11836153001) and PhosSTOP phosphatase (Roche, #4906845001) inhibitors. Lysates were centrifuged at 20,000g, 1h, 15°C, and the resulting supernatant was quantified by the Bradford assay (Bio-Rad, #5000201). To obtain phosphorylated fractions, above 400 µg of protein were separated for protein digestion. Proteins were reduced with DTT (final concentration of 20 mM; 30 min; room temperature), alkylated with iodoacetamide (final concentration of 30 mM; 30 min in dark; room temperature), diluted to 0.9 M with ABC, and digested with trypsin (Promega, #V5280) (1:20 w/w enzyme protein ratio, 18h, 37°C). Protein digestion was interrupted by acidification (acetic acid, pH<6), and the resulting peptides were cleaned up using Pierce Peptide Desalting Spin Columns (Thermo Fisher Sci., #89851). Phospho-peptide enrichment was performed using the High-Select TiO_2_ Phospho-peptide enrichment Kit (Thermo Fisher Sci., #A32993) according to the manufacturer’s instructions. Lastly, the enriched phospho-enriched fractions were cleaned up as described above and dried-down in a Speed Vacuum system. Aliquots of 10 µg cleaned up peptides from protein digestions were set aside for total protein analyses.

### Data independent acquisition (DIA) mass-spectrometry (MS)

Dried-down peptide samples were reconstituted with 2% ACN-0.1% FA (Acetonitrile-Formic acid), spiked with internal retention time peptide standards (iRT, Biognosys), and quantified by NanoDropTM spectrophometer (ThermoFisher Sci.) before LC-MS/MS in an EASY-1000 nanoLC system coupled to an EZ-Exploris 480 mass spectrometer (Thermo Fisher Sci.). Peptides were resolved using C18 Aurora column (75µm x 25cm, 1.6 µm particles; IonOpticks) at a flow rate of 300 nL/min using a 60-min gradient (50°C): 2% to 5% B in 1 min, 5% to 20% B in 48 min, 20% to 32% B in 12 min, and 32% to 95% B in 1 min (A = FA, 0.1%; B = 100% ACN:0.1% FA). Peptides were ionized using 1.6 kV spray voltage at a capillary temperature of 275 °C. We used data-independent acquisition (DIA) with full MS scans (scan range: 400 to 900 m/z; resolution: 60,000; maximum injection time: 22 ms; normalised AGC target: 300%) and 24 periodical MS/MS segments applying 20 Th isolation windows (0.5 Th overlap: Resolution: 15000; maximum injection time: 22 ms; normalised AGC target: 100%). Peptides were fragmented using a normalized HCD collision energy of 30%. MS data files were analysed using Spectronaut (Biognosys) by direct DIA analysis (dDIA). MS/MS spectra were searched against the Uniprot proteome reference from the *Homo sapiens* database <u>UP000005640</u> using standard settings. The enzyme was set to trypsin in a specific mode. On the one hand, Carbamidomethyl (C) was set as a fixed modification, and oxidation (M), acetyl (protein N-term), deamidation (N), and Gln to pyroGlu as variable modifications for total protein analysis. On the other hand, Carbamidomethyl (C) was set as a fixed modification, and oxidation (M), acetyl (protein N-term), and Phospho (STY) as variable modifications for phospho-proteome analysis. Identifications were filtered by a 1% Q-value. After MS, samples that did not pass QC were omitted from the study, resulting in a sample of G2019S L2PD (n=32) (15 from B and 17 from S), G2019S L2NMCs (n=22) (9 B and 13 S), R1441G L2PD (n=13) (1 B, and 12 D), R1441G L2NMCs (n=7) (2 B, and 5 D), iPD patients (n=39) (19 B, 10 S, and 10 D), and healthy controls (n=42) (23 B, 10 S, and 9 D). Lastly, to disambiguate peptide IDs into gene names we used the Uniprot online database (https://uniprotparser.proteo.info/).

### Proteome differential analysis

Proteome MS output data was exported from .SNE files from Spectronaut in a pivot table text format. For the differential analyses between groups, MS data was processed using QFeatures (doi: 10.18129/B9.bioc.QFeatures) in R (<u>QFeatures v1.13.1</u>). We applied the following R workflow: (i) Data was filtered to remove proteins identified by only 1 peptide sequence. (ii) Data selection was done based on condition and sub-group labels, with overall analysis containing all samples, G2019S analyses containing Barcelona and Santander samples labelled with prefix “B” or “S”, and R1441G analyses containing samples labelled with prefix “D” from Donostia-San Sebastian and “B” from Barcelona if bearing R1441G. For each analysis, we provided a separate Rscript file with a customized group selection (**Suppl. Material**). (iii) A protein ID column was assigned as an identification column for the analysis with QFeatures. (iv) We filtered out any row with 70% or more missing data. Here, with a 70% missing data cut-off, a meta-analysis would have a total of 3,815 rows while a more common 30% missing data cut-off would result in 3,789 rows. Since there was only about a 0.71% difference between the cut-off threshold, we chose the 70% cut-off so that we could keep entries potentially found in only one group without affecting the statistical power of the entire analysis. (v) Imputation of missing data was done using the kNN method (<u>QFeatures v1.13.1</u>). Subsequently, (vi) we performed a log2 transformation of the imputed data matrix, and (vii) designed a contrast matrix for differential analysis using limma.^36^ (viii) For each contrast matrix, we performed a limma analysis with false discovery rate (FDR) Benjamini & Hochberg (BH) multiple testing adjustment and collected the outputs under the criteria for statistical significance of an FDR-BH adjusted P<0.05 (1.12 log10) and a log_2_ fold-change (FC) above |0.6| (|1.5| in lineal values). Scripts for proteome raw data download and re-analysis are available online (**Suppl. Material**). For ANOVA analysis, we used the normalized data from above as starting point. The data from each row was grouped depending on the criteria used for grouping. Then for each comparison, we applied a Python script using one-way ANOVA analysis on the grouped data within the comparison and returned the P-value output as a new column.^37^ Then we perform the same FDR-BH correction from above to obtain the multiple testing adjusted P-values using the Statsmodels Python package^38^ with Python scripts also available online (**Suppl. Material**).

### Phospho-proteome differential analysis

Phospho-proteome MS data was exported from .SNE files from Spectronaut in a long-form table format using a Spectronaut param export file available online (**Suppl. Material**). Data was imputed using a modified version of a collapsing R script (<u>Perseus Plugin Peptide Collapse</u>)^39^ with phosphorylation as target modification at a confidence cut-off above 0.75. Modified collapsing.R and Perseus parameter.xml files are available online (**Suppl. Material**). We applied the following R workflow: (i) Columns with more than 70% blank cells were removed to meet the kNN requirement of less than 80% blank columns. (ii) Data selection for QFeatures input was based on condition and sub-group labels using all samples for overall analysis or specific group combination for location-specific and mutation-specific group combination, with overall analysis containing all samples, G2019S analyses containing Barcelona and Santander samples labelled with prefix “B” or “S”, and R1441G analyses containing samples labelled with prefix “D” from Donostia-San Sebastian and “B” from Barcelona when carrying R1441G. For each phospho-analysis group, we provide a separate Rscript file with customization to the selection group (**Suppl. Material**). Subsequently, (iii) we performed imputation by removing any row with 30% or more empty data similar to the proteome analysis using the kNN method, and (vi) performed log2 transformation normalization of the data using the quantile normalization method. (v) The statistical significance criteria were set at an FDR-BH adjusted P<0.05 (1.12 in log_10_) and a log_2_ fold-change (FC) above |0.6| (|1.5| in lineal values). (vii) In each differential analysis, we matched the protein and its original sequence using protein UniProt ID and performed the extraction of PTM position in protein and peptide, the peptide sequence, and the sequence window for visualization at the Curtain tool.^28^ Scripts for phospho-proteome data re-analysis are available online (**Suppl. Material**). For phospho-proteome ANOVA analysis, we followed the same methodology as for the proteome analysis but using the normalized phospho-proteome datasets from above. Data belonging to each group was identified from their column name. One-way ANOVA was applied on each row of cell groups from their respective comparison. The final statistically significant output values were adjusted using the Statsmodels package under the same FDR-BH multiple testing adjustment of P<0.05. Python scripts for ANOVA phospho-proteome analyses are also available online (**Suppl. Material**).

### Data visualisation

Aligning to FAIR principles^27^ of data findability, accessibility, interoperability, and reusability, we used Curtain and Curtain PTM,^28^ as free open-source tools for MS phospho-/proteomics data mining and exploitation by MS non-experts. Visualization of each of the differential analysis results from limma was done in volcano plot representation using the default cut-off settings of a fold-change (FC) above |1.5| (|0.6| log2) and a FDR-BH adj. P<0.05 (1.12 in log_10_). The Curtain tools enable interactively perusing volcano plots, deconvoluting primary experimental data to individual replicates that can be visualized in bar charts or violin plots allowing statistical analysis, and export of plots in .SVG format (<u>Curtain tutorials</u>). For each analysis, we also provide web links in the Figures and Figure legends. From each link, users can view the data associated with each data point on the volcano plot in the form of bar charts and violin plots. The magnitude of the data within these plots represents the relative intensity of the protein (total proteome) or phospho-site (phospho-proteome) before normalization. Beyond simple visualization of the numerical data, Curtain tools also aggregate data for different knowledgebases including UniProt, AlphaFold, PhosphoSitePlus, ProteomicsDB, and StringDB.

### Machine learning modelling of G2019S differential phospho-/proteins

The normalized and imputed datasets comprising differentially expressed peptides and phospho-peptides were employed to train a multi-class classifier to distinguish between Controls, G2019S L2PD and G2019S L2NMC. Three distinct candidate models were considered including Support Vector Machine (SVM), Random Forest (RF), and Gradient Boosting (GB) classifiers as described in other studies.^40^ Parameter optimization of the models through a grid search with a 5-fold cross-validation. To mitigate potential performance degradation due to unbalanced group sizes, we applied the Synthetic Minority Over-sampling Technique (SMOTE)^41^ to the training split. We used the balanced accuracy score^42^ defined as the average recall across each class, as a metric to evaluate models performances. Implementation of the models was done using the Scikit-learn^43^ v1.3.1 library within Python^44^ programming language v3.9.18.

### Classifier selection by comparative performance of machine learning models

In the G2019S proteome dataset, we included a total of 32 G2019S L2PD, 22 G2019S L2NMCs, and 42 controls that overpassed the QC criteria described above. Similarly, the phospho-proteome dataset comprised 29 G2019S L2PD, 19 G2019S L2NMCs, and 35 controls. First, we assessed comparative model performances for each dataset considering an initial number of features 3,816 peptides and 10,180 phospho-peptides respectively (**Suppl. Table 1**). Notably, in the proteome dataset, the SVM classifier demonstrated a substantial enhancement in balanced accuracy score following redundant feature elimination, achieving 0.91. This outcome indicates that the selective elimination of features contributed to obtaining a more discriminative model. Contrarily, the RF classifier showed limited improvement, implying that feature elimination methods were less effective for this specific model. Consistently, we obtained similar results for the phospho-proteome dataset where, after feature elimination, SVM achieved a balanced accuracy of 0.95, again highlighting the efficacy of feature selection in enhancing model performance. Furthermore, GB demonstrate significant improvement with only 43 features. This result indicates that the model performance can be enhanced with only a small subset of features. After comparative evaluation and parameter optimization, we identified SVM as the resulting most optimal model to derive informative LRKK2 signatures using the minimum subset of relevant features that maximize the discrimination between classes.

### Identification of a differential G2019S phospho-/protein signature

After SVM model selection, an initial set of relevant features was determined by incorporating only statistically significant features (P<0.05) identified by ANOVA test. Subsequently, we applied Recursive Feature Elimination with Cross-Validation (RFECV)^45^ to iteratively reduce the number of features while maximizing the balanced accuracy score. To obtain the LRKK2 signature, we employed the Monte Carlo Tree Search (MCTS)^46^ method. The MCTS strategy involved selecting the minimum combination of features that maximize the score in additive manner. Considering that the combinatorial features scale rapidly, the depth of the tree was fixed to five to manage computational complexity. The reward at each node of the tree was computed as the balanced accuracy score obtained through model training with cross-validation, utilizing the selected subset of features. At each iteration, the number of trees evaluated was set to 10 times the number of features. Following the evaluation of all the trees, the MCTS identified the best feature to add, maximizing the reward. A stop node was introduced to halt the algorithm when no further improvement could be achieved. In summary, the procedure comprised: (i) selection of the first feature, (ii) MTCS evaluation of all possible trees and reward calculation, (iii) selection of the best feature to be added, (iv) iteration from the second step until the model stops, (v) repetition from the first step until all features were screened. After the screening of all features, we selected the combinations of features with a balanced accuracy score above 0.90. The most prominently represented features were used as initial features for refinement by MCTS. Discriminant LRKK2 signatures were defined as the subset with the highest score after the refinement. Feature selection and refinement were implemented in Python v3.9.18 using Scikit-learn v1.3.1 and MCTS v2.0.4 libraries (https://pypi.org/project/monte-carlo-tree-search).

### Phospho-/protein gene ontology enrichment

Differential phospho-/proteins gene ontology (GO) was assessed using Metascape^47^ cell component term using default settings (min. overlap: 3, min. enrichment: |1.5|, P<0.05), and false discovery rate (FDR) adjusted P<0.05. Specifically, for signature phospho-/proteins, we used a combination of cell component and biological processes, and KEEGs, Reactome and wiki pathways under the same statistical significance cut-off.

### pSer106 RAB12 immunoblotting of 1-year follow-up PBMCs

To assess the levels of pSer106 RAB12 and other markers over time we isolated PBMCs using 40 ml of blood from a subset of subjects from Clínic-Barcelona (n=48), collected >1-year after DIA-MS, including G2019S L2PD (n=12), G2019S L2NMCs (n=6), iPD (n=15), and controls (n=15). Cell lysates were mixed with the NuPAGE LDS Sample Buffer (Life Technologies) supplemented 5% (v/v) 2-mercaptoethanol and heated to 90°C for 10 minutes. 10 μg of protein was loaded onto NuPAGE 4–12% Bis-Tris Midi Gels (Thermo Fisher, #WG1403BX10). Following the manufacturer’s instructions, samples were electrophoresed in NuPAGE MOPs SDS running buffer (Thermo Fisher). Protein transfer was performed onto nitrocellulose membranes (GE Healthcare, Amersham Protran Supported 0.45 μm NC) at 100 V for 90 min on ice in transfer buffer (48 mM Tris–HCl and 39 mM glycine supplemented with 20% methanol) or through turbotransfer. MLi-2 treated protein extracts were electrophoresed in 4–15% Mini-PROTEAN® TGX™ Precast Protein Gels (Bio-Rad, #4561084) and transfer onto trans-Blot Turbo Mini 0.2 µm Nitrocellulose Transfer (BIORAD, #1704158) through a Trans-Blot Turbo Transfer System (BIORAD) following the manufacturer’s instructions. Following transfer, membranes were blocked using 5 (w/v) skim milk dissolved in TBS-T (20 mM Tris–HCl, pH 7.5, 150 mM NaCl, and 0.1% (v/v) Tween 20) at RT, for 30 minutes. Membranes were then washed once with TBS-T before they were incubated overnight at 4°C with primary antibodies diluted in 5% BSA (Sigma) dissolved in TBS-T. Membranes were washed 3x with TBS-T for 5 min each before incubation with secondary antibodies diluted in TBS-T at RT for 1 hour. Following secondary incubation, the membranes were then washed 3x with TBS-T for 10 min each. Acquisition of protein bands was performed with near-infrared fluorescent detection using the Odyssey CLx imaging system and quantified using the Image Studio software. The sheep monoclonal anti-total RAB12 antibody was purified by MRC PPU Reagents and Services at the University of Dundee and was used at a final concentration of 1 μg/ml. Mouse monoclonal anti-GAPDH (Santa Cruz Biotechnology, sc-32233) was used at 1:10,000. In-house generated mouse monoclonal anti-total LRRK2, rabbit monoclonal anti-pSer935 LRRK2 (Abcam, #ab133450), rabbit polyclonal anti-total RAB12 (Protein Tech, #18843-1-AP), rabbit monoclonal anti-pSer106 RAB12 (Abcam, #ab256487), rabbit monoclonal anti-RAB9A (Cell Signalling Technologies, #5118), rabbit monoclonal anti-LAMP1 (Cell Signalling Technologies, #9091), and mouse monoclonal anti-Tubulin (Cell signalling Technologies, #3873S) were used at a 1:1,000 dilution (**Suppl. Table 2**). Secondary antibodies, anti-mouse (Licor, #926-68022), anti-rabbit (Licor, #926-32213), and anti-goat (Licor, #926-68074), which has sheep cross-reactivity, were used at a 1:10,000 dilution.

### pSer106 RAB12 immunoblotting after MLi-2 LRRK2 inhibition in fresh PBMCs

To assess the response of pSer106 RAB12 to MLi-2 LRRK2 inhibition we additionally isolated PBMCs from 40 ml of peripheral blood of an additional set of subjects (n=10), including G2019S L2PD (n=3), R1441G L2PD (n=1), iPD (n=1), and healthy controls (n=5). These cells were collected > 2 years after DIA-MS. Each 20 ml of freshly extracted PBMCs was aliquoted into two technical replicates of two tubes of 5 ml each to perform MLi-2 pharmacological inhibition following the Dundee PBMCs isolation protocol (doi: <u>dx.doi.org/10.17504/protocols.io.bnhxmb7n</u>) (from whole blood). Briefly, each technical replicate was treated with either 200 nM MLi-2 LRRK2 inhibitor or an equivalent volume of DMSO (5 µl) for 30 min at room temperature. Next, treated PBMCs were centrifuged at 355g for 5 min. Each pellet was resuspended in 1 ml of PBS containing 2% FBS with or without 200 nM MLi-2, and transferred into an Eppendorf tube. After centrifugation at 300 g for 3 min, treated PBMC pellets were lysed with 70 µl of ice-cold lysis buffer (50 mM Tris–HCl, pH 7.5, 1% Triton X-100, 1 mM EGTA, 1 mM sodium orthovanadate, 50 mM NaF, 0.1% 2-mercaptoethanol, 10 mM 2-glycerophosphate, 5 mM sodium pyrophosphate, 0.1 µg/ml mycrocystin-LR, 270 mM sucrose, 0.5 mM DIFP and Complete EDTA-free protease inhibitor cocktail) on ice for 10min. Then, treated PBMC cell lysates were centrifuged at 14,000 rpm 15 min at 4°C and supernatant was collected. Protein concentrations were assessed at a 1:15 dilution through the Bradford assay. Protein extracts were immediately used for immunoblotting or snap-frozen and stored at −80°C. Subsequent immunoblot analysis was done as described above, with some specificities: Brielfly, MLi-2 treated protein extracts were electrophoresed in 4–15% Mini-PROTEAN® TGX™ Precast Protein Gels (Bio-Rad, #4561084) and transferred onto trans-Blot Turbo Mini 0.2 µm Nitrocellulose Transfer (Bio-Rad, #1704158) through a Trans-Blot Turbo Transfer System (Bio-Rad) following the manufacturer’s instructions. Acquisition of protein bands for MLi-2 protein extracts was performed on LAS4000 system and processed through the BandPeak plugin (doi: <u>dx.doi.org/10.17504/protocols.io.7vghn3w</u>) at the ImageJ software. For this experiment, mouse monoclonal anti-RAB10 (Merck, #SAB5300028), rabbit recombinant anti-pThr73 RAB10 (Abcam, #ab230261), rabbit polyclonal anti-RAB12 (Protein Tech, #18843-1-AP), rabbit monoclonal anti-p106 RAB12 (Abcam, #ab256487) were used at a 1:1,000 dilution (**Suppl. Table 2**). As secondary antibodies we used goat anti-rabbit (ThermoFisher, #31460) and goat anti-mouse (Abcam, #ab205719).

### Clinical correlation of LRRK2 differential phospho-/proteins and disease severity

We performed a Spearman’s association analysis between the differential proteins and phospho-proteins across different comparisons (log2FC>|0.6|, adj. P<0.05) and UPDRS-III motor scores from PD patients and healthy controls. To this end, we used the “cor.test” function from R (stats v4.3.1) to calculate Rho coefficients and the EnhancedVolcano package (v1.20.0) to represent correlation outputs. Statistical significance was set a Spearman’s correlation coefficient Rho>|0.5| and an FDR multiple-testing adjusted P<0.05.

## RESULTS

We succeeded in quantifying the levels of 3,798 unique proteins using DIA-MS from a LRRK2 clinical cohort (n=174) (**Fig. 1**). Pairwise analysis, under a cut-off of ≥2 peptide mapping, <0.30 imputation, log_2_FC>|0.60|, and adj. P<0.05, revealed that G2019S L2PD was the most distinct group displaying a set of 207 proteins whose levels differed vs controls, with 85% down-regulated proteins (168 down/ 39 up) (**Fig. 2**). We found that in G2019S L2PD a number of proteins expression was reduced namely, ATIC, which can repress LRRK2 and rescue neurodegeneration^48^ (log_2_FC=-0.97, adj. P=1.92×10^-13^); RAB9A, involved in phagocytic vesicle trafficking and lysosomal function (log_2_FC=-1.17, adj. P=3.97×10^-10^); or LAMP1, a lysosome biogenesis and autophagy regulator (log_2_FC=-1.32, adj. P=1.63×10^-9^). G2019S L2NMCs vs controls showed 67 differential hits, involving mostly down-regulated proteins (57 down/ 10 up), common and with the same FC direction as L2PD (42 of 67), e.g., ATIC or LAMP1 (**Suppl. Fig. 1**). G2019S L2PD vs L2NMCs differed only in 2 proteins down-regulated in G2019S L2PD, RAB9A (log_2_FC=0.77, adj. P=0.038) and SCLY, a Selenocysteine lyase involved in peptide elongation (log_2_FC=1.58, adj. P=0.038). These results indicate proteome changes common to all G2019S carriers and associated with G2019S.

**Fig. 1.**
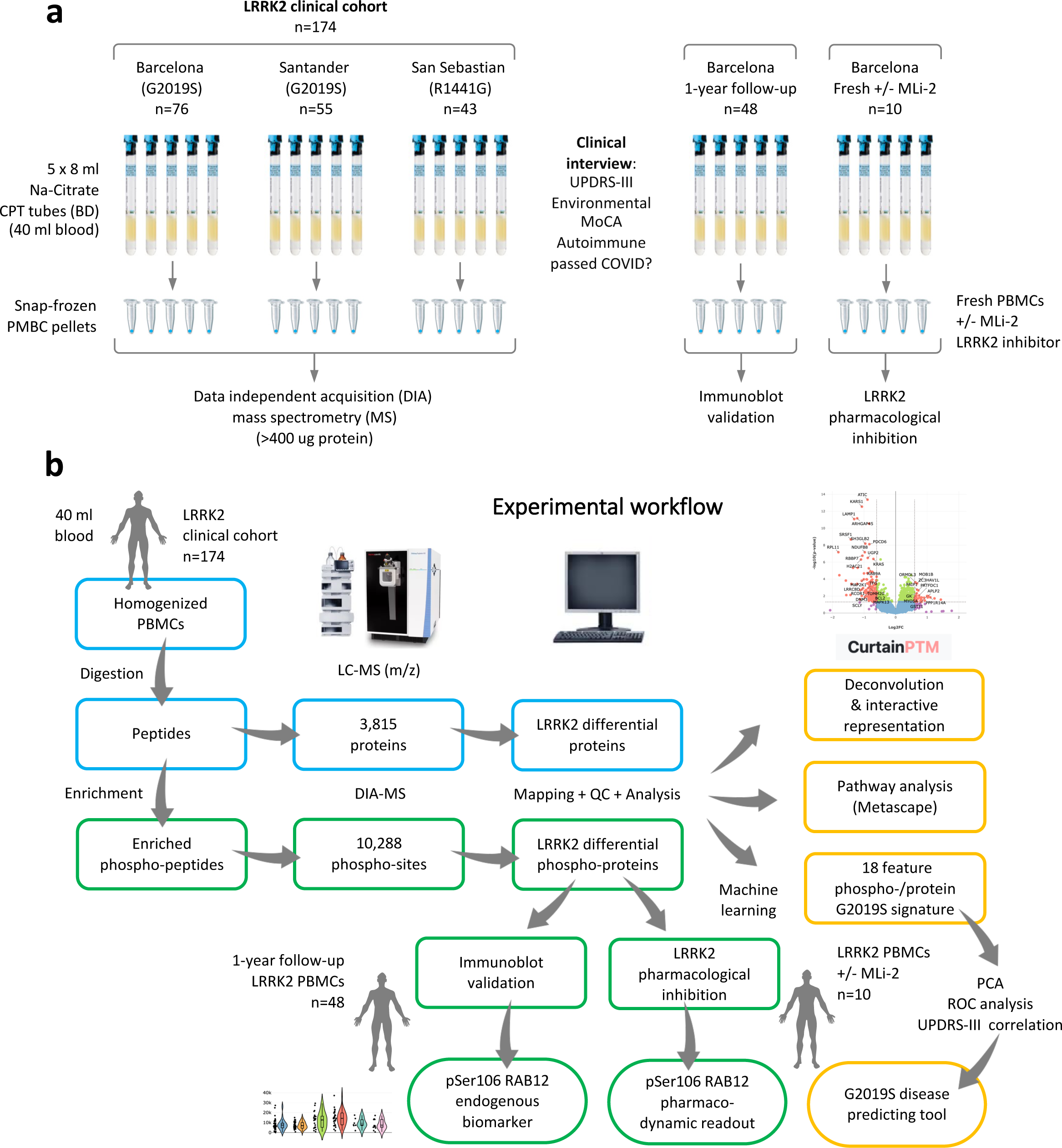
Experimental workflow using PBMCs from a Spanish LRRK2 clinical cohort. (**a**) Peripheral blood mononuclear cells (PBMCs) processing for different applications. 40 ml of blood were drawn from subjects of a LRRK2 clinical cohort from Spain (n=174) encompassing G2019S L2PD patients (n=37), G2019S L2NMCs (n=27), R1441G L2PD patients (n=14), R1441G L2NMCs (n=11), iPD (n=40), and controls (n=45). (**b**) After PBMCs isolation, homogenization, and protein digestion, a total of 3,815 proteins were identified by DIA-MS on an EZ-Exploris 480 mass-spectrometer (Thermo), and 10,288 phospho-sites after phospho-enrichment. For the group differential analysis, we only considered proteins and phospho-sites mapped by ≥2 different peptides (Spetronaut), and with <30% imputation, with a significance cut-off of log_2_FC>|0.6| and a multiple testing adjusted P<0.05. Data deconvolution and interactive representation of findings were done using the Curtain / Curtain PTM Tool, and gene ontology was assessed by Metascape. Using machine learning, we identified an 18-feature G2019S phospho-/protein signature able to discriminate G2019S L2PD, G2019S L2NMCs, and controls. By immunoblot, we assessed pSer106 RAB12 / total RAB12 levels in PBMCs from a subject of subjects (n=48) after 1 year of follow-up, including G2019S L2PD (n=12), G2019S L2NMCs (n=6), iPD (n=15) and controls (n=15). Lastly, in freshly isolated PBMCs from a second subset of subjects (n=10) encompassing G2019S L2PD (n=3), R1441G L2PD (n=1), iPD (n=1) and healthy controls (n=5), treated with DMSO or the MLi-2 LRRK2 inhibitor, we performed an LRRK2 kinase assay measuring pSer106 RAB12 / total RAB12 levels.

**Fig. 2.**
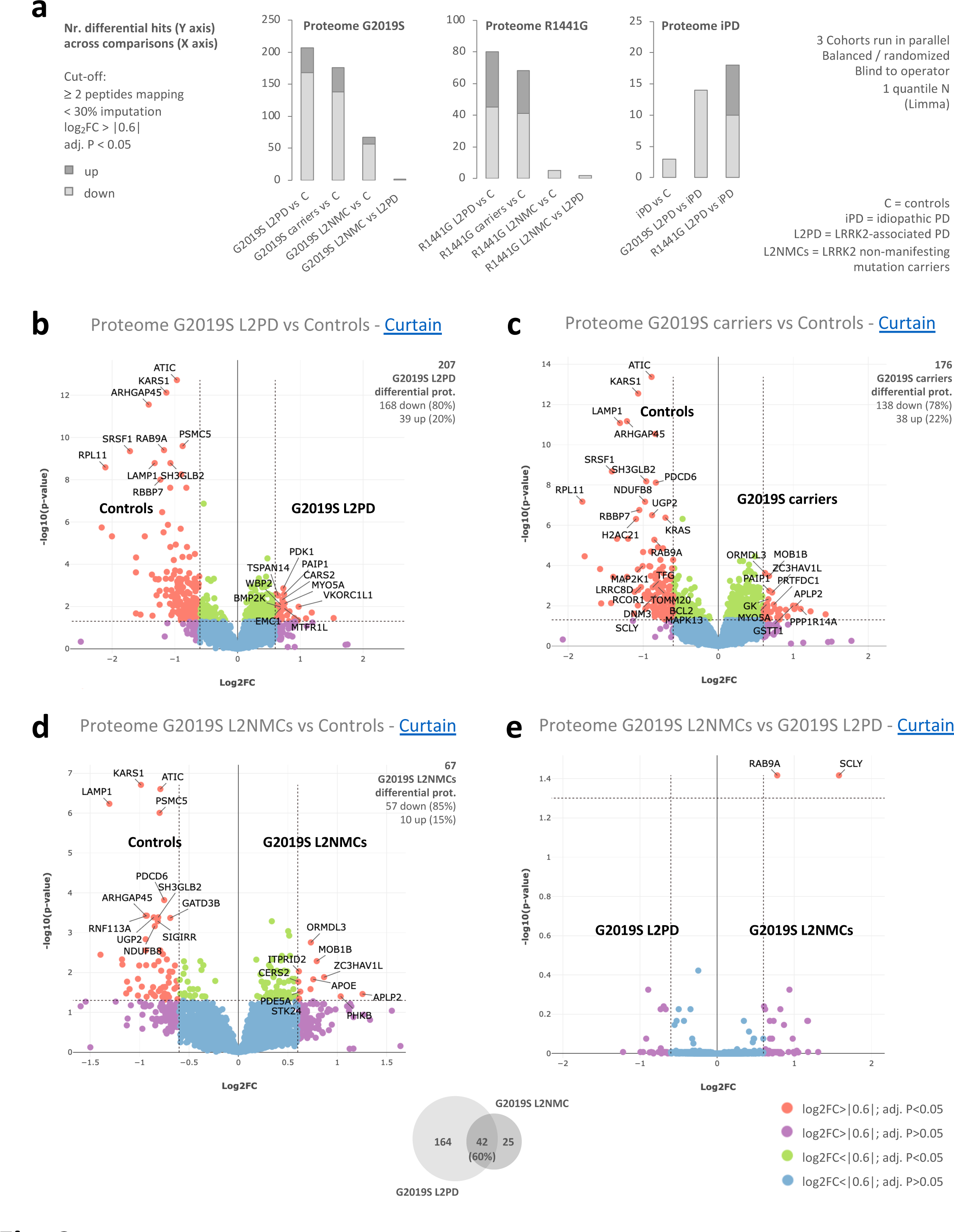
Proteome overview and differential analyses in G2019S carriers. (**a**) Barplots showing the numbers of differential proteins in different pairwise comparisons involving G2019S carriers, R1441G carriers, iPD, and controls, with up-regulated proteins in dark grey, and down-regulated in light grey. All cohorts were run in parallel, with balanced study groups per run, blind to the operator, and using 1 quantile normalisation (Limma). The significance cut-off was set at a log_2_FC>|0.6| and a multiple testing adjusted P<0.05. (**b**) Volcano plot of the proteome differential analysis in G2019S L2PD vs healthy controls, with Curtain weblinks to access raw and differential analysis data, showing proteins up-regulated in G2019S L2PD as red dots on the right, and proteins up-regulated in controls (i.e., down-regulated in G2019S L2PD) as red dots on the left (<u>Curtain</u>). A legend colour code applying to all panels is shown at the bottom of the figure, depicting statistically significant hits as red dots. (**c**) Volcano plot of the proteome differential analysis in G2019S carriers as a whole, i.e., L2PD and L2NMCs, vs healthy controls (<u>Curtain</u>). (**d**) Volcano plot showing the proteome differential analysis between G2019S L2NMCs and healthy controls (<u>Curtain</u>). (**e**) Volcano plot representing the proteome comparison between G2019S L2NMCs and G2019S L2PD. A Venn diagram at the bottom of the figure shows the overlap of differential hits in PD-manifesting and non-manifesting G2019S carriers (<u>Curtain</u>).

Regarding the R1441G proteome, R1441G L2PD vs controls revealed 80 hits (45 down/ 35 up) (**Suppl. Fig. 2**). Of these, 33 proteins (30 down/ 3 up) (44%) overlapped with G2019S L2PD and had the same FC direction, including down-regulation of NDUFB8, a mitochondrial Complex I subunit; PDCD6, a calcium sensor involved vesicle trafficking and apoptosis; RPL11, a component of the 60S ribosomal subunit; and hits such as ATIC, RAB9A, LAMP1, and SLCY. R1441G L2NMCs vs controls showed 5 down-regulated proteins, all common to R1441G L2PD, including NDUFB8 and PDCD6. Between R1441G L2PD and L2NMCs, 2 proteins were up-regulated in R1441G L2PD; ATG3, an E2 ubiquitin-like conjugating enzyme; and MAGT2, which is essential for Golgi protein N-glycosylation. iPD vs controls, despite their larger sample, had only 3 differential hits, all down-regulated and common to L2PD, i.e., SRSF1, an RNA splicing factor; UQCRB, a mitochondrial Complex III subunit; and LAMP1 (**Suppl. Fig. 1 and 4**). Such findings might be related to the clinical heterogeneity of iPD due to diverse genetic and environmental factors. Functionally, proteome changes in G2019S and R1441G L2PD, even iPD, revealed a shared biological enrichment affecting endolysosomal trafficking, proteostasis, and mitochondrial function (**Suppl. Fig. 3**).

Regarding the G2019S phospho-proteome, we found 10,288 phospho-sites mapping to 2,657 proteins in PBMCs. Using the same stringent cut-off from above, G2019S L2PD vs controls displayed a single differential phospho-site, pSer106 RAB12, hyper-phosphorylated in G2019S L2PD (log_2_FC=0.97; adj. P=0.036) and L2NMCs (log2FC=0.92; adj. P=0.057) (**Fig. 3**). Remarkably, pSer106 RAB12 was shown as a key physiological LRRK2 substrate of higher expression than other RABs in brain from PD models, including pThr73 RAB10.^49,50^ G2019S carriers as a whole showed elevated levels of pSer106 RAB12 (log_2_FC=0.95; adj. P=0.003) and pTyr334 SKAP2 (log_2_FC=1.05; adj. P=0.003), a protein involved in immune response at peripheral tissues which regulates neural functions at the CNS,^51^ including a-synuclein phosphorylation.^52^ G2019S L2NMCs vs L2PD showed increased pSer205 MON2 (log_2_FC=1.25; adj. P=0.05), a regulator of endosome to Golgi trafficking. Lastly, we found no differential hit of G2019S L2NMCs compared to controls. Collectively, these results identify enhanced pSer106 RAB12 levels in a large cohort of G2019S carriers, pinpointing for the first time pSer106 RAB12 as an endogenous biomarker in G2019S PBMCs.

**Fig. 3.**
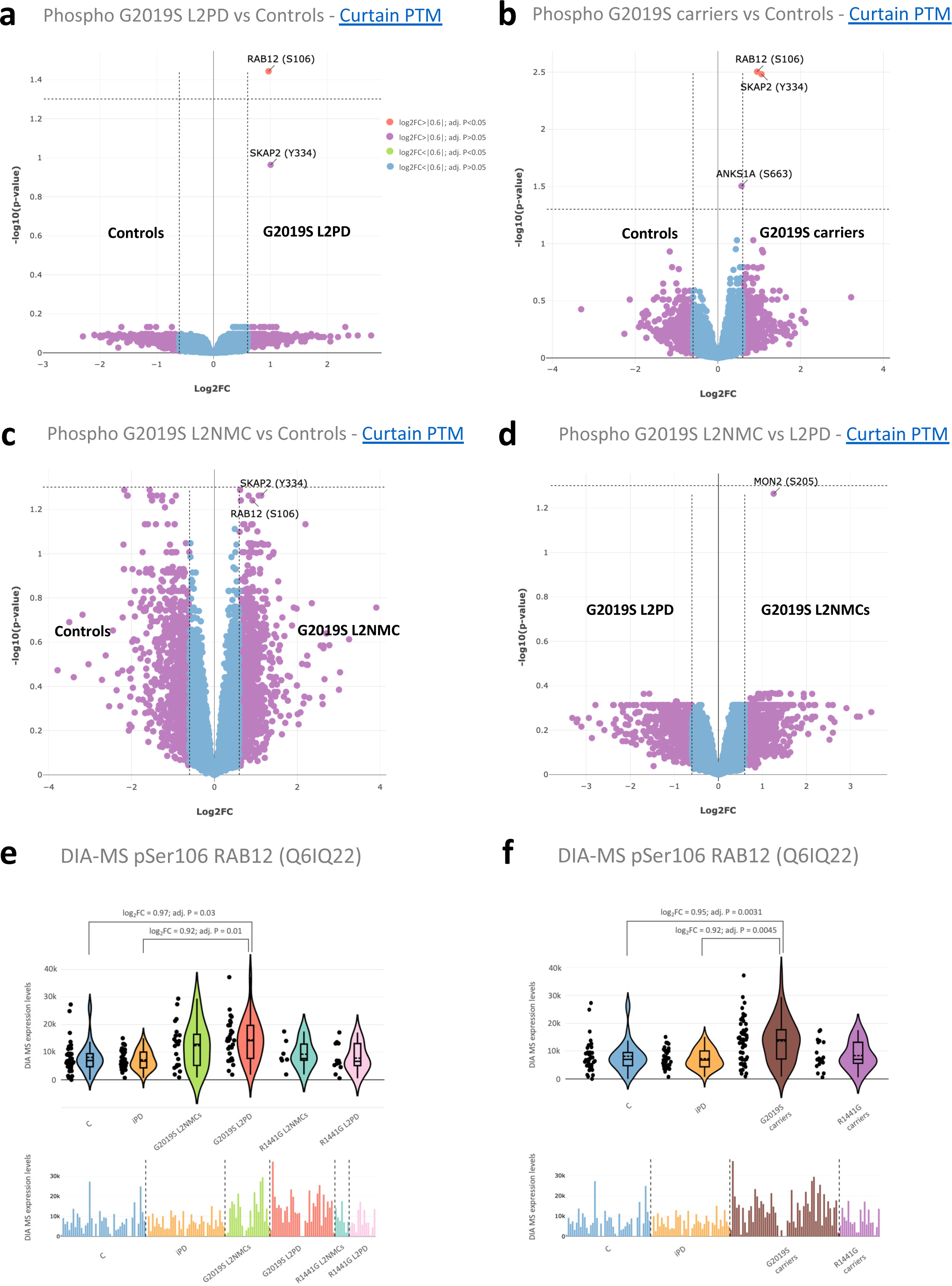
Phospho-proteome differential analyses of G2019S carriers. (**a**) Volcano plot of the phospho-proteome differential analysis of G2019S L2PD vs controls, and Curtain weblinks to raw and differential analysis data, showing hyper-phosphorylated proteins in G2019S L2PD as red dots on the right, where a single hit, elevated pSer106 RAB12 levels in G2019S L2PD, emerged (<u>Curtain PTM</u>). A legend colour code shows hits categorisation by statistical significance and applies to all the panels. (**b**) Volcano plot showing phospho-protein hits in G2019S carriers as a whole, PD-manifesting and non-manifesting, compared to controls (<u>Curtain PTM</u>). (**c**) Phospho-proteome differences in G2019S L2NMCs vs controls (<u>Curtain PTM</u>). (**d**) Volcano showing phospho-proteome differences in G2019S L2NMCs vs G2019S L2PD (<u>Curtain</u> <u>PTM</u>). (**e**) QC crude non-imputed (lower bar plot), non-normalised (upper violin plot) mass-spectrometry data from pSer106 RAB12 levels across all study groups showing higher pSer106 phosphorylation levels in G2019S L2PD and G2019S L2NMCs respect to the rest of the groups. The adj. P-values and FC on top of the violin plot correspond to those from the differential analysis. (**f**) A similar analysis to the previous panel with G2019S L2PD and G2019S L2NMCs grouped into a single group of G2019S carriers.

By phospho-proteome analysis of R1441G carriers, R1441G L2PD vs controls showed no hit overpassing the multiple-test adjustment. In addition, R1441G L2NMCs vs controls had 25 differential phospho-sites (20 down/ 5 up), but none included pSer106 RAB12 (**Suppl. Fig. 2**). These findings indicate that enhancement of pSer106 RAB12 phosphorylation is a specific effect in G2019S PBMCs, and suggest distinct phospho-signalling preferences occurring for different pathogenic LRRK2 mutations in G2019S and R1441G patients. Regarding iPD, at the phospho-proteome level, we found no phospho-site change compared to controls (**Suppl. Fig. 4**). However, iPD revealed large phospho-site differences to G2019S L2PD (84 down/ 9 up), including pSer106 RAB12, whose levels were elevated in G2019S L2PD, and also to R1441G L2PD (409 down/ 225 up). Altogether these findings indicate that phospho-protein changes are more prominent in L2PD due to phospho-signalling dysfunction by LRRK2 activating mutations than in iPD, being pSer106 RAB12 a preferred LRRK2 substrate in G2019S PBMCs rather than R1441G or iPD.

By immunoblot we assessed pSer106 RAB12 levels as pSer106 RAB12 / Total RAB12 ratios using >1-year follow-up PBMCs of the G2019S cohort from Clínic-Barcelona (n=48), encompassing G2019S L2PD (n=12), G2019S L2NMCs (n=6), iPD (n=15), and controls (n=15) (**Table 2**). Consistent with DIA-MS data, we found phosphorylation differences across groups (Kruskal-Wallis P=0.01), with borderline increased pSer106 RAB12 phosphorylation levels in G2019S L2PD (Dunn’s adj. P=0.069) and L2NMCs (Dunn’s adj. P=0.118) vs controls, and in G2019S carriers as a whole vs controls (Kruskal-Wallis P=0.003; Dunn’s adj. P=0.027), but not in iPD (**Fig. 4**, **Suppl. Fig. 5**). By immunoblot^53^ we did not observe down-regulation of proteome hits such as RAB9A / GAPDH in G2019S L2PD or iPD (Kruskal-Wallis P=0.08) nor LAMP1 / GAPDH except in iPD (Kruskal-Wallis P=0.03) (Dunn’s adj. P=0.046). Lastly, we assessed pSer106 RAB12 response to LRRK2 pharmacological inhibition by Mli-2 in technical replicates from freshly collected PBMCs of an additional set (n=10) of 3 G2019S L2PD, 1 R1441G L2PD, 1 iPD, and 5 controls, treated with Mli-2 (200 nM; 30 min) or DMSO (**Suppl. Fig. 6**). In all subjects, we observed a strong diminishment of pSer106 RAB12 phosphorylation levels after MLi-2 treatment, confirming pSer106 RAB12 as a pharmaco-dynamic readout of LRRK2 inhibition using human PBMCs.

**Fig. 4.**
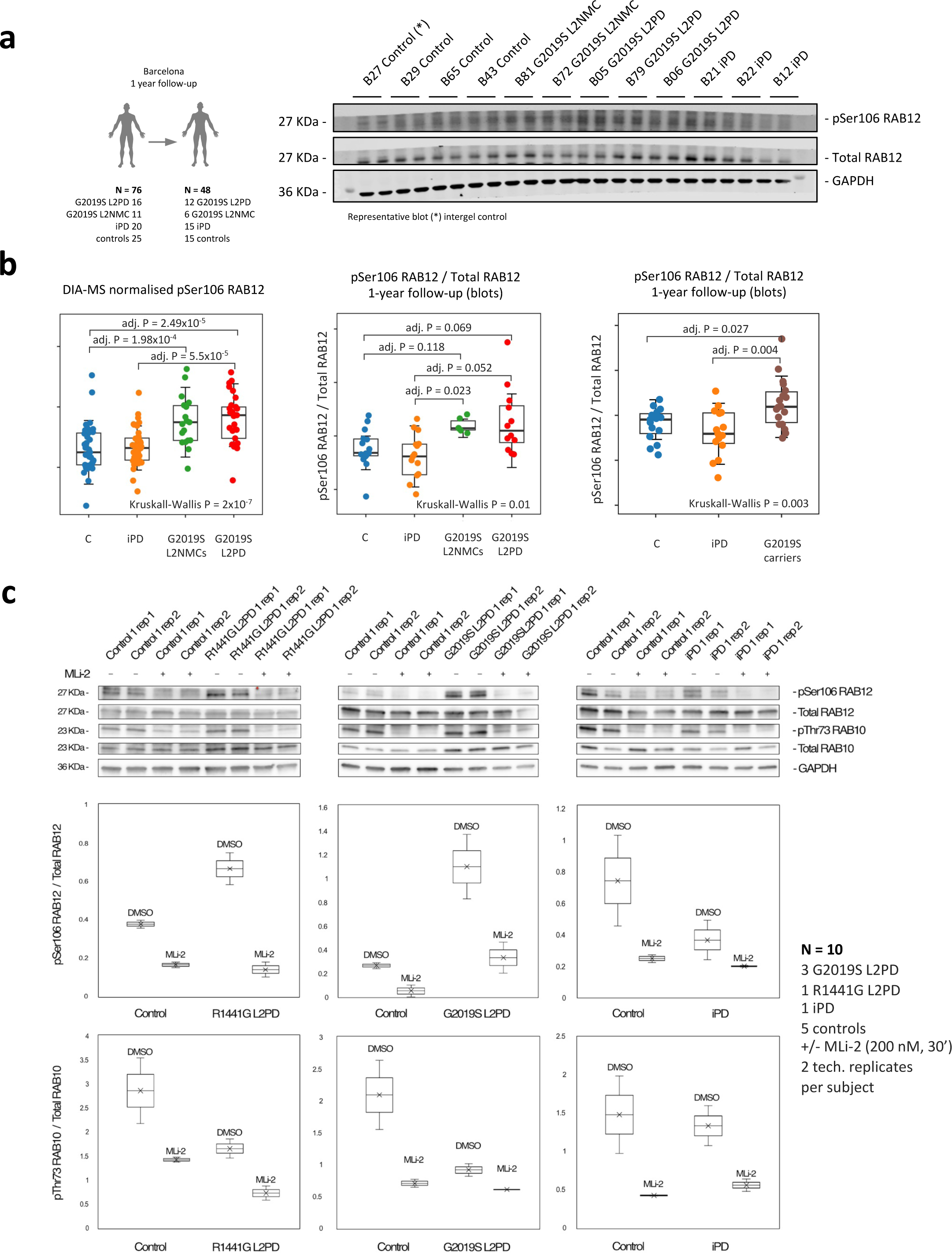
One-year follow-up of pSer106 RAB12 by immunoblot and MLi-2 response. Immunoblot assessment of pSer106 RAB12 phosphorylation levels in >1-year follow-up PBMC samples from part of the LRRK2 subcohort from Clínic-Barcelona (n=48), including G2019S L2PD (n=12), G2019S L2NMCs (n=6), iPD (n=15), and controls (n=15). (**a**) Schematic workflow of immunoblot assessment and representative blot from 5 different blots shown in the Supplement. (*) Denotes intergel control. (**b**) dot plots comparing pSer106 RAB12 / Total RAB12 levels obtained by DIA-MS at the entire LRRK2 clinical cohort (n=174) on the left, and by immunoblot of part of the Clínic-Barcelona cohort after 1-year of follow-up (n=48) in G2019S carriers on the right. In each plot, overall intergroup differences were assessed using the Kruskal-Wallis test followed by post-hoc Dunn’s test to assess for pSer106 RAB12 / Total RAB12 differences in G2019S carriers. (**c**) Representative immunoblot analysis of pSer106 RAB12 / Total RAB12 and pThr73 RAB10 / Total RAB10 using technical replicates from additional freshly collected PBMCs from one R1441G L2PD, one G2019S L2PD, one iPD and 3 controls (expanded to a total n=10 subjects in the Supplement), treated with DMSO or the MLi-2 LRRK2 inhibitor (200 nM, 30 min), showing a diminishment of pSer106 RAB12 phosphorylation levels after LRRK2 inhibition by MLi-2 treatment.

Next, we interrogated phospho-/protein subsets able to classify G2019S carriers and controls. We applied a supported vector machine (SVM) classifier, adjusting for unbalanced group sizes, with 5-fold cross-validation as overfitting control. After recursive feature elimination, we found 510 peptides and 204 phospho-sites subjected as multi-class informative items. By Montecarlo Tree Search (MCTS), we refined combinations of minimal numbers of features yielding maximal balanced accuracy. We found an 18-feature G2019S signature of 15 proteins and 3 phospho-sites (**Fig. 5**), including pSer106 RAB12 among others, e.g., ATIC, RAB9A, LAMP1, NDUFB8, SCLY, or pSer205 MON2, that provided a balanced accuracy to discriminate groups of 0.96 and an area under the curve (AUC) of 1.00 for G2019S L2PD vs controls, and of 0.99 for G2019S L2NMCs. Despite being developed upon G2019S data, the classifier also discriminated R1441G carriers from controls, but not iPD, indicating some common features to R1441G. The top gene ontology term of the 18 features was vesicle transport. We also found a 17-feature G2019S signature involving only proteins (**Suppl. Fig. 7**). These results indicate biological plausibility and 96% disease-prediction ability of the 18-feature phospho-/protein signature to classify G2019S carriers by both mutation and disease status.

**Fig. 5.**
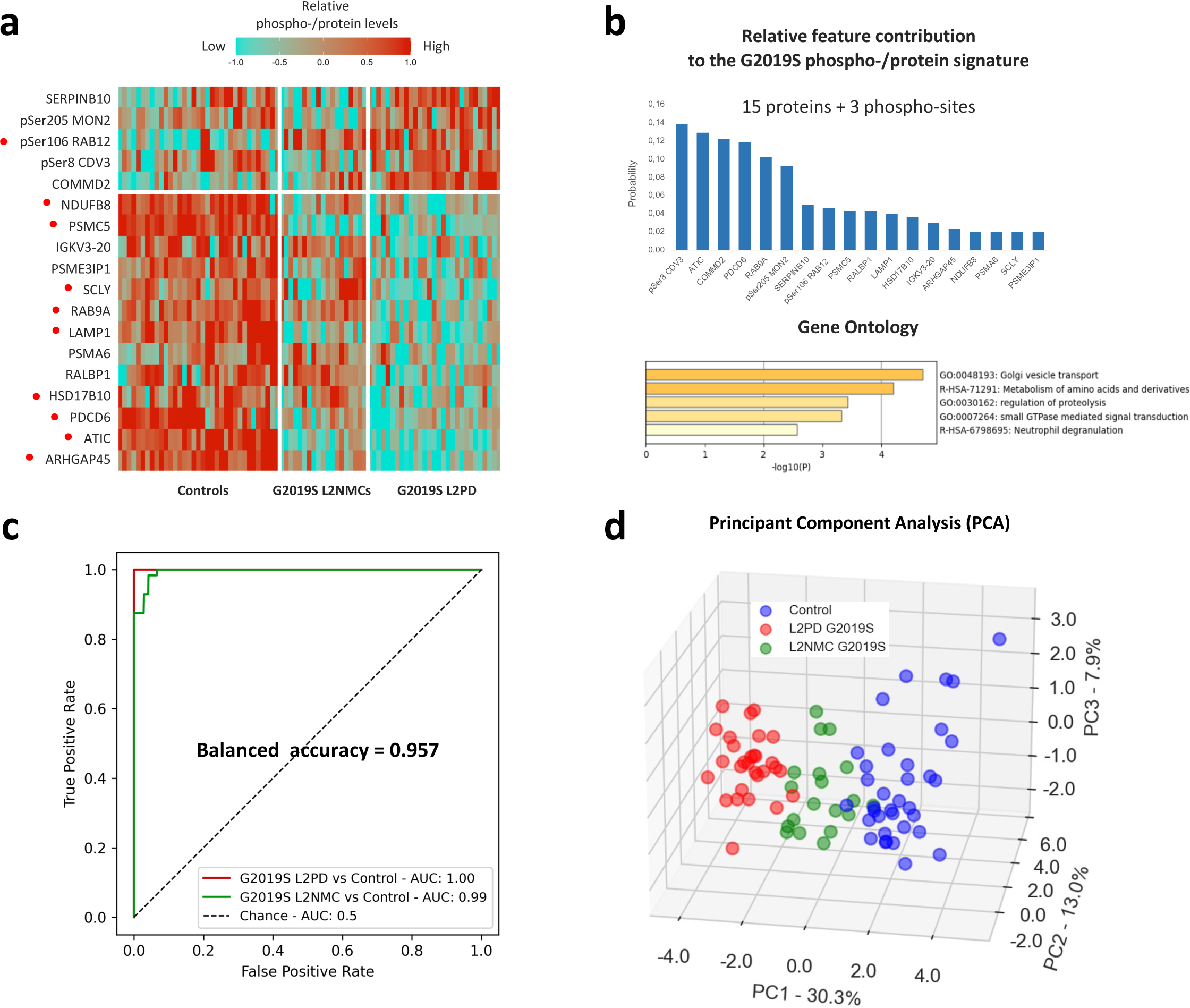
Identification of an 18-feature phospho-/protein classifier for G2019S carriers. After comparing the performance of several models, we applied supported vector machine (SVM) learning, adjusted by unbalanced groups using the Synthetic Minority Over-sampling Technique (SMOTE), corrected from overfitting with 5-fold cross-validation, identified cross-group differential proteins and phospho-proteins by ANOVA and Recursive Feature Elimination with Cross-Validation (RFECV), and refined informative combinations to the minimal numbers of features yielding the maximal balanced accuracy by the Montecarlo Tree Search (MCTS) method. (**a**) 18-feature G2019S phospho-/protein best classifier identified in G2019S carriers, both PD-manifesting and non-manifesting subjects, and healthy controls. Red dots indicate individual features correlating with disease severity (UPDRS-III) (See next Figure). (**b**) Relative contribution of the different proteins (n=15) and phospho-sites (n=3), including pSer106 RAB12, from the 18-feature G2019S classifier on the upper bar plot; Metascape gene ontology enrichment analysis of the 18-features G2019S signature lower bar plot. (**c**) Receiver Operating Curve (ROC) analysis of the 18-feature G2019S phospho-/protein signature showing an overall balanced accuracy of 0.957 to discriminate G2019S L2PD, G2019S L2NMCs and controls, specifically with an area under the curve (AUC) of 1.00 between G2019S L2PD and controls, and 0.99 between G2019S L2NMCs and controls. (**d**) Principal component analysis (PCA) based on the 18-feature G2019S phospho-/protein classifier in G2019S carriers and healthy controls showing distinct group profiles based on LRRK2 mutation and disease status, with G2019S L2NMCs in between G2019S L2PD and controls, consistent with their disease status.

Lastly, we interrogated whether deregulated phospho-/proteins were related to disease severity, as assessed by UPDRS-III motor scores. Under a Spearman’s Rho>|0.5| and a P<0.05, we found 34 differential proteins (16%) of the 207 hits in G2019S L2PD vs controls with inverse association with UPDRS-III whereas pSer106 RAB12 and pSer205 MON2 had a direct correlation with motor scoring. Moreover, 10 markers (55%) from the 18-feature G2019S phospho-/protein correlated inversely with UPDRS-III (ATIC, PDCD6, RAB9A, PSMC5, LAMP1, HSD13B10, ARHGAP45, NDUFB8, and SCLY), meanwhile pSer106 RAB12 correlated positively (Rho = 0.49, adj. P = 1.60×10^-4^) (**Fig. 6**). PDCD6, the top common correlating protein (Rho=-0.75, P=5.51×10^-10^), participates in vesicle trafficking, mediates mitochondrial cytochrome c release and apoptosis,^54^ and has been linked to PD.^55^ We also found 65 out of the 80 hits (81%) in R1441G L2PD vs controls correlating with UPDRS-III, both inverse (59%) and positively (41%), several of which were present at the 18-feature G2019S signature (PDCD6, ARHGAP45, NDUFB8, RAB9A, ATIC, SCLY, and LAMP1) whereas others were exclusive for R1441G, e.g., the mitochondrial protein UBQLN4 (Rho=-0.89, P=1.64×10^-6^) or the cytoskeletal protein PLEC (Rho=0.84, P=3.50×10^-5^). Although correlation does not mean causality, these results indicate that some phospho-/proteins at the 18-feature G2019S classifier can be related to disease severity therefore holding potential clinical relevance.

**Fig. 6.**
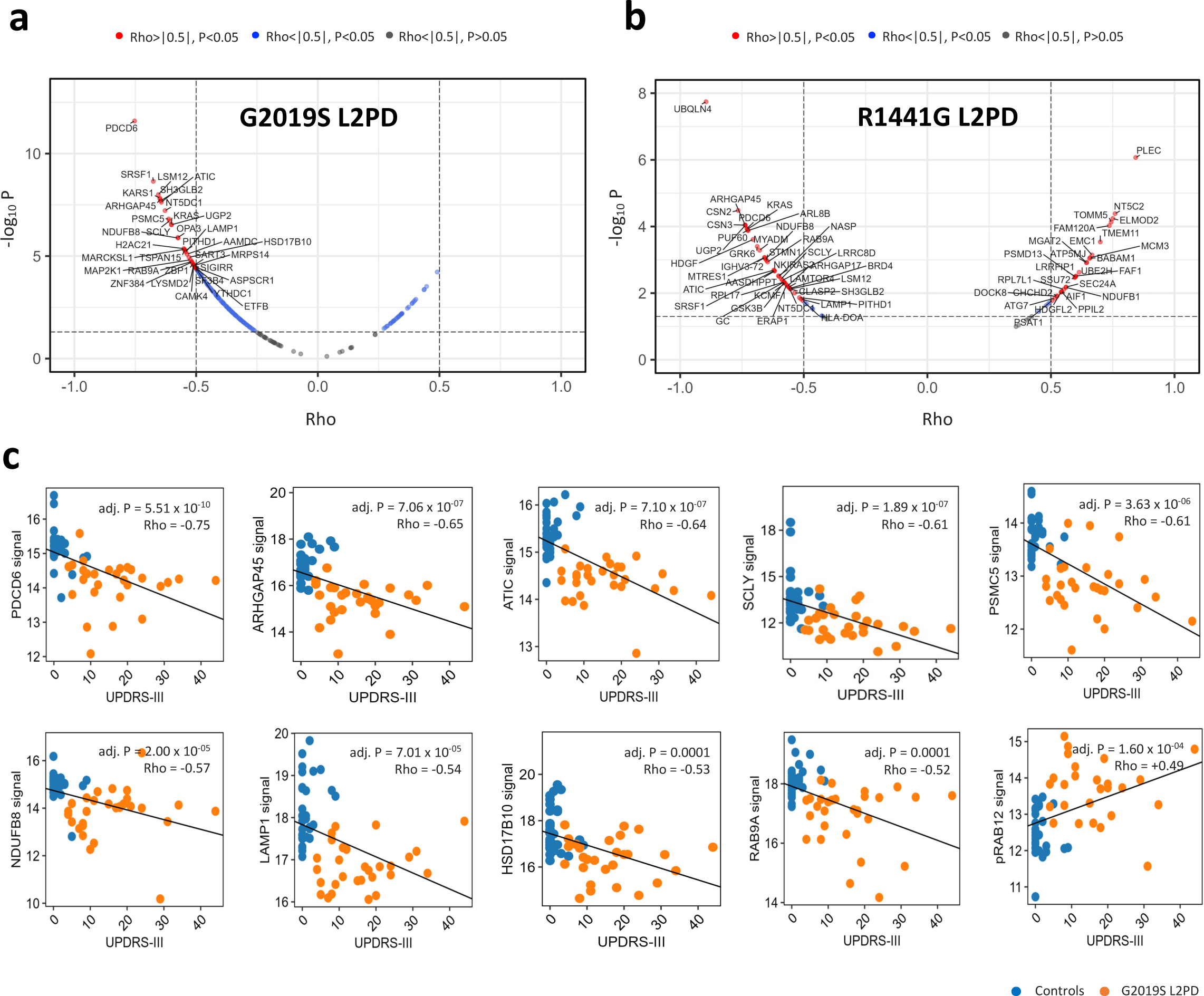
Association between differential LRRK2 phospho-/proteins and disease severity. Correlation analysis of differential proteins and phospho-proteins (log2FC>|0.6|, adj. P<0.05) and UPDRS-III motor scores from L2PD patients and healthy controls with statistical significance set at a Spearman’s correlation coefficient Rho>|0.5| and an FDR multiple-testing adj. P<0.05. (**a**) Correlation plots between differential proteins in G2019S L2PD vs controls on the left, and R1441G L2PD vs controls on the right, showing differential hits correlating with UPDRS-III in red. (**b**) Scatter plot of 10 hits from the 18-feature G2019S phospho-/protein signature correlating with UPDRS-III in G2019S L2PD patients represented as orange dots and healthy controls as blue dots, including PDCD6, ARHGAP45, ATIC, SCLY, PSMC5, NDUFB8, LAMP1, HSD17B10, RAB9A, and pSer106 RAB12.

## DISCUSSION

Following FAIR principles,^27^ we employed an interactive tool called Curtain^28^ in which the raw and differential analysis data is saved with weblinks which, can be readily explored by non-MS experts. The G2019S L2PD proteome, showed the highest number of changes, approx., 200 proteins, most of which were down-regulated (80%). The G2019S L2NMCs displayed fewer protein differences, around 70, which were also downregulated (85%). There was a strong overlap between proteins that changed in both groups (60%). Comparing G2019S L2PD and G2019S L2NMCs revealed two proteins (RAB9A and SCLY) enhanced in the asymptomatic carriers. Our findings indicate prominent protein deficits mostly associated with LRRK2 mutations such as G2019S, which can begin at G2019S L2NMCs premotor stages^56^ and progress in G2019S L2PD.

Metascape gene ontology analysis annotated the protein changes in the G2019S carriers as participating in the endolysosomal biology cycle, involving vesicle trafficking and mitochondrial function. For example, G2019S L2PD showed down-regulation of RAB9A, which controls phagocytosis and lysosomal biology.^57,58^ In G2019S carriers we also observed down-regulated levels of LAMP1, a canonical lysosomal marker involved in lysosome biogenesis which supports enhanced LRRK2 activity.^7–9^ A previous study also noted that LAMP1 levels were reduced in CSF of L2PD.^59^ Our findings are consistent with the current understanding of the LRRK2 pathway indicating that it plays a key role in controlling the endosomal lysosomal pathway.^50,60^

Beyond endolysosomal changes, we also observed changes in proteins involved in ribosomal function, protein homeostasis, and alternative splicing related to G2019S.^61^ Thus, ATIC, the top protein down-regulated in G2019S carriers, catalyses the last two steps of mitochondria purine biosynthesis^62,63^ and was also linked previously to LRRK2 toxicity.^48^ Other deficits included KARS1, a tRNA synthetase; PSMC5, the proteasomal 26S subunit; or SCLY, seleno-cysteine lyase, an enzyme involved in peptide elongation that is also involved in neurodegeneration.^64^ Our findings in LRRK2 PMBCs align with studies reporting transcriptional repression of proteostasis regulators in G2019S L2PD,^65^ and proteostasis defects in PD substantia nigra.^66^

R1441G L2PD showed 80 differential proteins with 40% shared with G2019S L2PD. Enrichment analysis showed that the functions of the proteins deregulated in the R1441G carriers were similar to G2019S. The R1441G L2PD top protein deficit, NDUFB8, is a subunit of the mitochondrial Complex I (NADH to Ubiquinone oxidoreductase), whose activity is deficient in PD.^67^ R1441G L2PD and L2NMCs displayed few protein differences, notably ATG3, involved in autophagy, and MGAT2, a Golgi glycosyl transferase. Overall the proteomic effects of the G2019S and R1441G LRRK2 mutations in our clinical cohort were quite similar.^68,69^

The iPD proteome, despite being the largest group, displayed only 3 differential hits that were commonly decreased in G2019S and R1441G L2PD. These were LAMP1, which further supports endolysosomal dysfunction occurring in iPD;^60,70^ SRSF1, a Serine/Arginine-rich splicing factor; and UQCRB, a mitochondrial Complex III subunit (Ubiquinol-cytochrome c oxidoreductase). Beyond the etiopathological heterogeneity of iPD,^71,72^ the relatively fewer protein changes in iPD than G2019S or R1441G L2PD suggest more specific effects of the LRRK2 mutations in dysregulating signal transduction pathways in LRRK2 mutants than in iPD.

Regarding the phospho-proteome, a single hit, pSer106 RAB12, was found to be specifically elevated in the G2019S but not R1441G carriers. Excitingly this phospho-site comprises a key physiological substrate of LRRK2.^2^ Overall the roles that RAB12 plays and its phosphorylation by LRRK2 are poorly understood. Phosphorylation of RAB12 is prominent in the brain and observed higher than other RAB substrates such as RAB10 in this organ.^49,50^ Other studies showed that RAB12 is located in phagosomes, late endosomes, and lysosomes where it may regulate endosome to trans-Golgi trafficking and exocytosis.^73,74^ To our knowledge, this is the first report of hyper-phosphorylated RAB12 in PBMCs from a large cohort of G2019S carriers.

We analysed n=48 follow-up PBMC samples after 1 year by immunoblotting. Despite the lower sample, we found an increase of pSer106 RAB12 in G2019S L2PD and L2NMCs. Previous studies in neutrophils probing for RAB10 but not RAB12 phosphorylation revealed elevated pThr73 RAB10 in R1441G but not G2019S carriers.^19^ We also found no RAB10 phosphorylation increase in G2019S and R1441G PBMCs by DIA-MS. Such results in G2019S PBMCs suggest that either RAB12 is a preferred substrate for LRRK2 - indeed, distinct mutation effects cannot be ruled out,^75^ and/or that pThr73 RAB10 phosphatases, e.g., PPM1H,^76^ dephosphorylate RAB10 more efficient than RAB12. Mechanistic studies on how G2019S and other LRRK2 variants preferentially phosphorylate different RABs in various cell types, using larger cohorts, are warranted. Our study identifies pSer106 RAB12 as an endogenous biomarker in easily accessible PBMCs from carriers of the most prevalent G2019S mutation, either L2PD or L2NMCs.

Upstream of LRRK2, PD cell models showed LRRK2 activation by VPS35/ RAB29 (RAB7L1) binding to a region on the Armadillo (ARM) domain termed ‘Site-1’.^22,23^ More recently, RAB12 was shown as a key LRRK2 activator that binds to a distinct site at the ARM domain termed ‘Site-3’.^24,25^ One study showed that RAB12 played a role in recruiting LRRK2 to damaged or stressed lysosomes.^25^ These studies suggested that ARM domain Site-1 or Site-3 inhibitors that block RAB binding could serve as novel therapeutic target for allosteric inhibitors of LRRK2 kinase activity.^24^ The biological effect of pSer106 RAB12 phosphorylation on LRRK2 regulation has not been well characterised and our results emphasize that additional work is warranted to investigate this.

Downstream of LRRK2, MLi-2 phospho-proteomics identified RAB3A, RAB8A, RAB10, RAB12, RAB29, and RAB43 as LRRK2 substrates.^2,3,20,77^ In the clinical setting, only pThr73 RAB10 has been validated as an LRRK2 substrate^19^ and exploited as a readout of LRRK2 activity in previous studies,^18^ including in LRRK2 inhibitor clinical trials.^12,21^ As mentioned above, there has not been a specific way of assessing elevated LRRK2 activity in G2019S carriers due to the lack of effect in pThr73 RAB10 phosphorylation. Monitoring pSer106 RAB12 phosphorylation levels could be especially useful for assessing G2019S selective inhibitors that have been newly developed in clinical studies,^78–81^ as these would be expected to preferentially reduce pSer106 RAB12 phosphorylation in patients with heterozygous G2019S mutations.

For G2019S carriers we also identified a signature of 15 proteins and 3 phospho-sites including pSer106 RAB12 that was found to provide a 96% accuracy to discriminate G2019S L2PD, L2NMCs, and controls. Although correlation does not imply causality, signature features such as ATIC, PDCD6, RAB9A, PSMC5, LAMP1, HSD13B10, ARHGAP45, NDUFB8, SCLY, pSer2015 MON2, and pSer106 RAB12 correlated with PD motor severity (UPDRS-III) suggesting that the phospho-signature may be related to PD progression, but further work with larger number of cohorts would be required to assess this clinically.^56^ This is the first signature in G2019S PBMCs based on DIA-MS data and complements previous G2019S signatures in blood^82^ and urine.^83,84^

Despite the exciting findings, our study has limitations. Inherent variation in humans markedly affects differential protein expression and phosphorylation. Slightly different procedures in PBMCs preparation and storage at different centres can also affect results. To minimise this variation, we undertook MS analysis and data analysis at the same time with blind groups. Third, differential enrichment of phospho-peptides on titanium dioxide beads can result in further variety. Indeed, we discarded one of the phospho-peptide batches due to not passing quality control. Due to the phospho-peptide enrichment approach used, the detection of phospho-Tyrosines was under-represented. We used a stringent significance cut-off filtering in only hits mapped by at least 2 peptides, and we cannot rule out that other important proteins could have been excluded. Lastly, the number of R1441G carriers, especially L2NMCs, was significantly smaller than G2019S and it was insufficient to assess signatures by machine learning.

In summary, aligning with urine,^84^ in PBMCs we found elevated pSer106 RAB12 levels as an endogenous biomarker for G2019S carriers. Given that RAB12 was shown as a key LRRK2 activator in PD models able to increase pThr73 RAB10 levels,^24,25^ future studies ought to investigate the effect of pSer106 RAB12 phosphorylation on LRRK2 activation. In line with PD models,^50,85^ in human LRRK2 PBMCs we found pSer106 RAB12 as a pharmaco-dynamic readout of MLi-2 LRRK2 inhibition. We also found an 18-feature signature including pSer106 RAB12 with high accuracy in discriminating symptomatic, asymptomatic G2019S carriers, and controls. Future studies need to assess pSer106 RAB12 in other G2019S clinical cohorts, blood cells and CSF, using precise quantitative methods such as reaction monitoring (RM) and ELISA-like assays, to hopefully translate our findings for clinical trials of novel LRRK2 inhibitors.^78–81^

## PUBLICLY AVAILABLE DATA

The mass spectrometry proteomics data have been deposited to the ProteomeXchange Consortium via the PRIDE^86^ partner repository with the dataset identifiers PXD050865 for the proteome and PXD050944 phospho-proteome analyses. Following FAIR principles,^27^ we through the interactive tool called Curtain,^28^ raw and differential analysis data is also provided as weblinks which to be readily explored by non-MS experts. Beyond that, programming scripts for data analyses (**Suppl. Material**) (to be provided).

## FUNDING

(Please insert only direct funding contributing to this study)

This study was supported by the Michael J. Fox Foundation for Parkinson’s Research (MJFF) (#MJFF-000858) to RFS, ME, CM, ES, JI, and JRM.

## Data Availability

We provide full open access to all data generated here through the information included in this article and the open Curtain software.

https://curtain.proteo.info/#/ce36322f-b1ef-4ee6-b454-6ffd5c8a0429

https://curtain.proteo.info/#/e97b6761-dbe3-41cb-8b3d-4ab9f2c93aba

## ACKNOWLEDGEMENTS

We thank the patients and their relatives for their continued and essential collaboration. LDM was funded by the Beatriu-de-Pinòs programme (#BP00176) from the Agència de Gestió d’Ajuts Universitaris i de Recerca (AGAUR). AR was funded by the PFIS programme (#FI21/00104) from the Instituto de Salud Carlos III (ISCIII) co-funded by the European Union. MF was funded by the María-de-Maeztu programme (#MDM-2017-0729) to the Parkinson’s disease and Movement Disorders group of the Institut de Neurociències (Universitat de Barcelona). ARC was funded by the EU Next-Generation 2022 Investigo programme from the European Commission (EC) / Agència de Gestió d’Ajuts Universitaris i de Recerca (AGAUR). Research in the Alessi lab were funded by UK Medical Research Council (grant number MC_UU_00018/1) and the pharmaceutical companies supporting the Division of Signal Transduction Therapy Unit (Boehringer Ingelheim, GlaxoSmithKline, and Merck KGaA). RFS was supported by a Miguel Servet grant (#CP19/00048), a FIS grant (#PI20/00659), a PFIS grant (#FI21/00104), and a M-AES grant (#MV22/00041) from the Instituto de Salud Carlos III (ISCIII) co-funded by the European Union. IDIBAPS receives support from the CERCA program of Generalitat de Catalunya.

## CONFLICTS OF INTEREST

None of the authors declare conflict of interest.

## FINANCIAL DISCLOSURE

None of the authors declare conflict of interest.

---

**Suppl. Table 1.** Detail of the performance of each model across (phospho-)/proteome datasets.

| Proteome |  |  |  |  |
| --- | --- | --- | --- | --- |
| Model | Initial<br>(3,816 features) | ANOVA<br>(926 features) | ANOVA +<br>RFECV | Feature<br>optimal Nr. |
| SVM | 0.69 ± 0.09 | 0.85 ± 0.08 | 0.91 ± 0.07 | 510 |
| RF | 0.68 ± 0.09 | 0.70 ± 0.12 | 0.67 ± 0.08 | 536 |
| GB | 0.69 ± 0.08 | 0.82 ± 0.13 | 0.67 ± 0.09 | 881 |
| Phospho-proteome |  |  |  |  |
| Model | Initial<br>(10,180 features) | ANOVA<br>(979 features) | ANOVA +<br>RFECV | Feature<br>optimal Nr. |
| SVM | 0.39 ± 0.11 | 0.79 ± 0.03 | 0.95 ± 0.07 | 204 |
| RF | 0.41 ± 0.10 | 0.53 ± 0.03 | 0.61 ± 0.10 | 91 |
| GB | 0.40 ± 0.08 | 0.47 ± 0.07 | 0.81 ± 0.06 | 43 |

SVM = Support Vector Machine; RF = Random Forest; GB = Gradient Boosting

**Suppl. Table 2.** Primary antibodies list.

| Antibody Description | Company | Cat. Number | Dilution / Concentration |
| --- | --- | --- | --- |
| Mouse monoclonal GAPDH (6C5) | Santa Cruz | #sc-32233 | 1:10,000 |
| Rabbit monoclonal LAMP1 (D2D11) | Cell Signalling Tech. | #9091 | 1:1,000 |
| Mouse monoclonal Total LRRK2 | In house generated | - | 1:1,000 |
| Rabbit pSer935 LRRK2 [UDD2 10(12)] | Abcam | #ab133450 | 1:1,000 |
| Rabbit monoclonal RAB9A (D52G8) | Cell Signalling Tech. | #5118 | 1:1,000 |
| Mouse monoclonal RAB10 | Merck | #SAB5300028 | 1:1,000 |
| Rabbit recombinant pThr73 RAB10 (MJF-R21) | Abcam | #ab230261 | 1:1,000 |
| Sheep polyclonal Total RAB12 | MRC-PPU Reagents and Services at the University of Dundee | #SA227 | 1 µg/ml |
| Rabbit monoclonal pSer106 RAB12 (MJF-R25-9) | Abcam | #ab256487 | 1:1,000 |
| Mouse monoclonal Tubulin (DM1A) | Cell Signalling Tech | #3873S | 1:1,000 |

**Suppl. Fig. 1.**
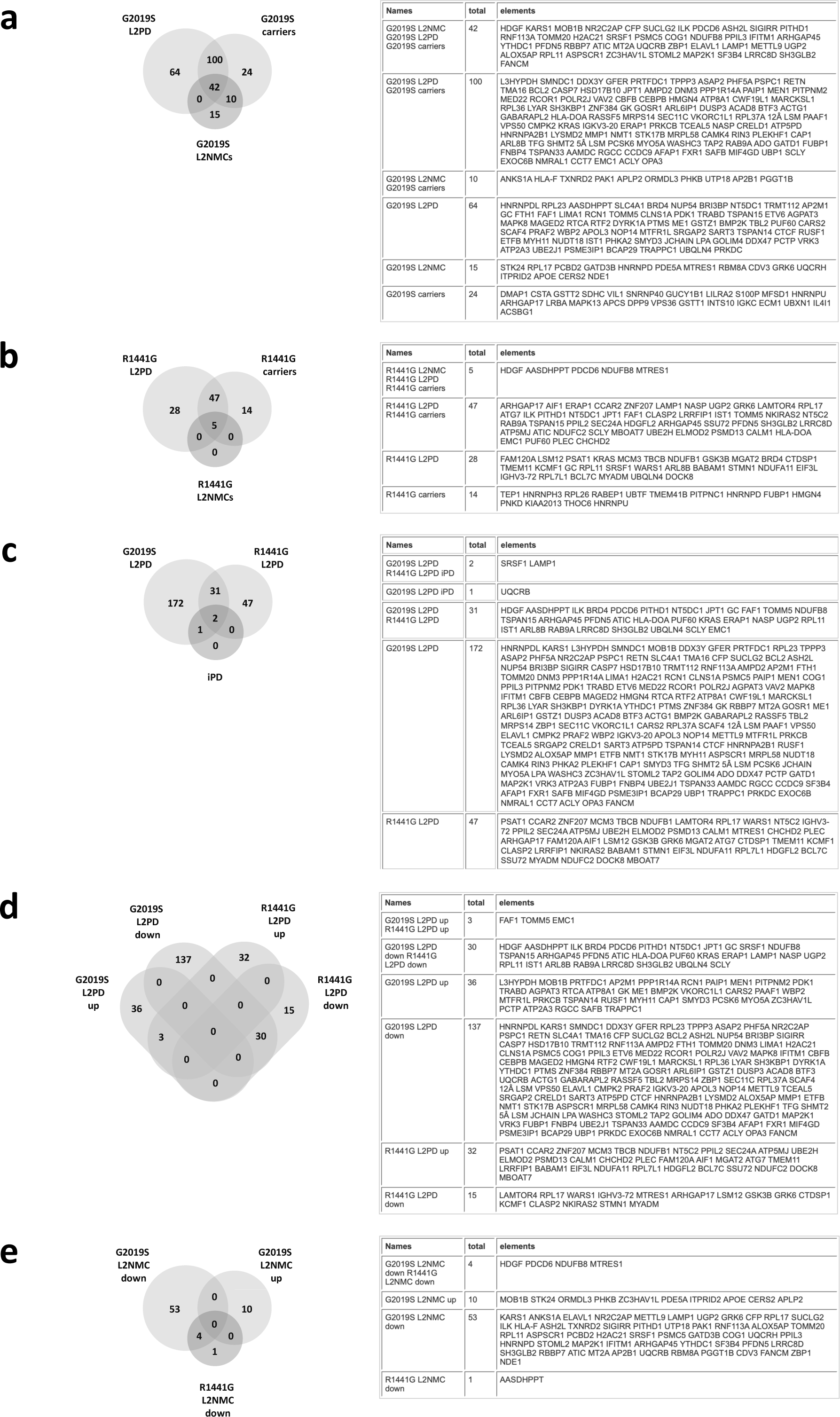
Comparison of proteome hits identified across different groups. Venn diagrams depicting common and specific hits in various groups as compared to healthy controls. (**a**) Differential hits found at various G2019S carrier groups, PD manifesting and non-manifesting, compared to controls. (**b**) Hits from various R1441G carrier groups, symptomatic and asymptomatic, vs controls. (**c**) Common and specific hits were observed in the different PD patient groups, i.e., G2019S L2PD, R1441G L2PD, and iPD. (**d**) Differential hits among L2PD patients carrying either the G2019S or the R1441G mutations, stratified by up and down-regulated hits. (**e**) Differential hits among L2NMCs carrying either the G2019S or the R1441G mutations, as analysed segregated by up and down-regulated hits.

**Suppl. Fig. 2.**
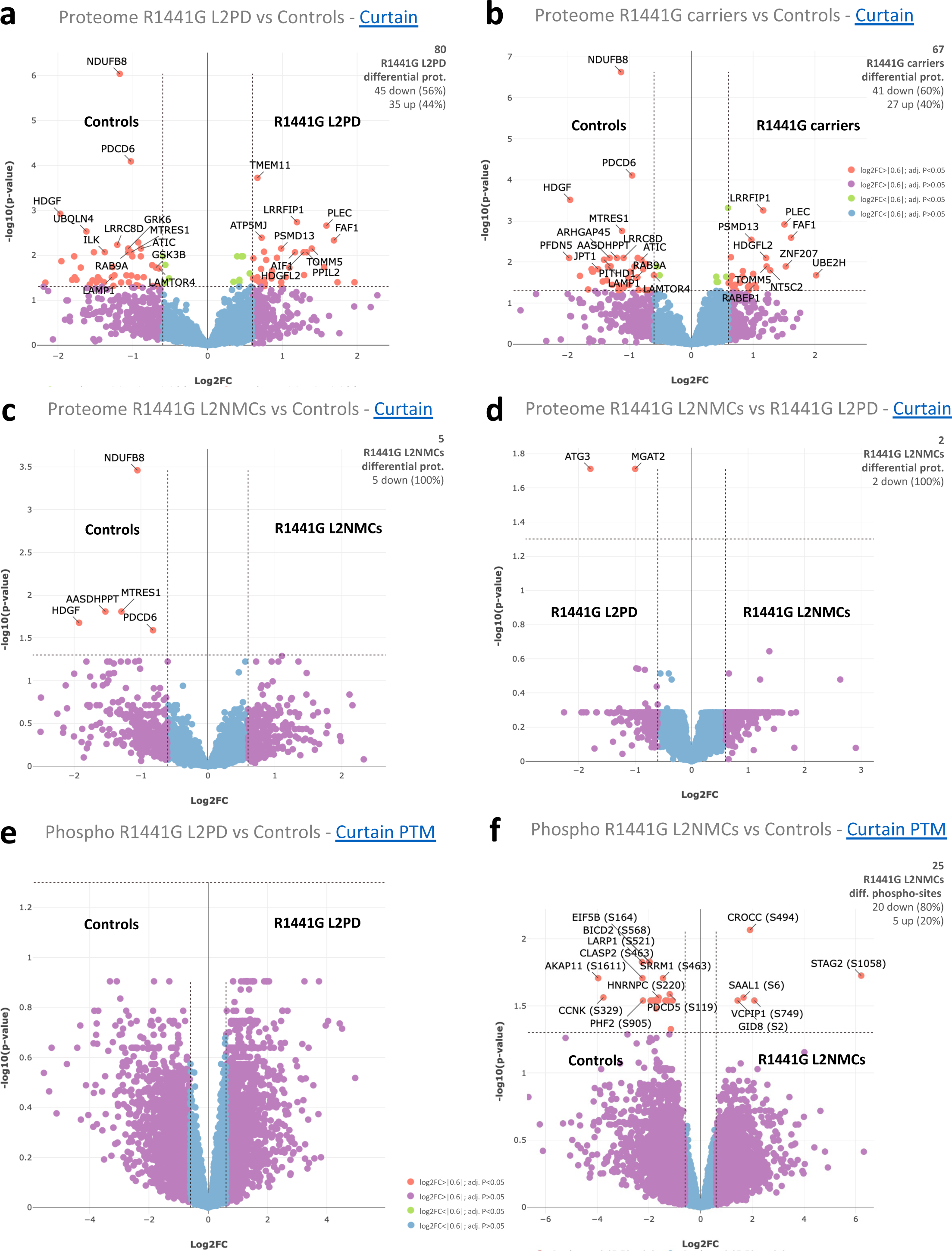
Proteome and phospho-proteome differential analysis of R1441G carriers. Consistently across the study, the significance cut-off for R1441G proteome and phospho- proteome analyses was also set at a log_2_FC>|0.6| and a multiple testing adj. P<0.05. A legend colour code applying to all panels is shown at the bottom of the figure, depicting statistically significant hits as red dots. (**a**) Volcano plot of the proteome differential analysis in R1441G L2PD vs healthy controls, with Curtain weblinks to access raw and differential analysis data, showing proteins up-regulated in R1441G L2PD as red dots on the right, and proteins up-regulated in controls (i.e., down-regulated in R1441G L2PD) as red dots on the left (<u>Curtain</u>). (**b**) Volcano plot of the proteome differential analysis in R1441G carriers as a whole, i.e., L2PD and L2NMCs, vs healthy controls (<u>Curtain</u>). (**c**) Volcano plot showing the proteome differential analysis between R1441G L2NMCs and healthy controls (<u>Curtain</u>). (**d**) Volcano plot representing the proteome comparison between R1441G L2NMCs and R1441G L2PD (<u>Curtain</u>). (**e**) Volcano plot of the phospho-proteome differential analysis between R1441G L2PD vs controls (<u>Curtain PTM</u>). (**f**) Volcano plot of the phospho-proteome comparison of R1441G L2NMCs and controls (<u>Curtain</u> <u>PTM</u>).

**Suppl. Fig. 3.**
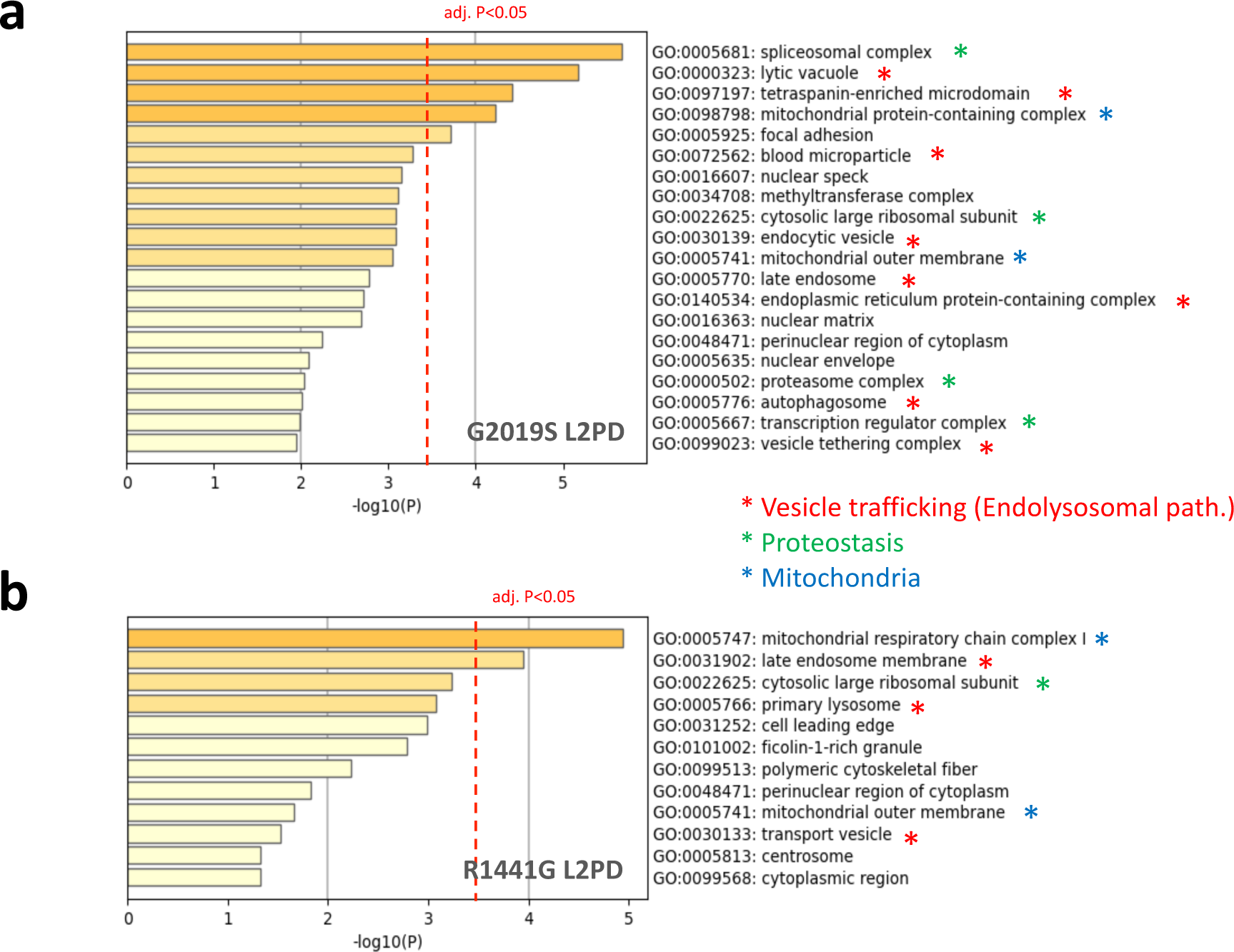
Proteome functional analysis of G2019S and R1441G patients. Comparative gene ontology (GO) enrichment analysis of the differential proteins observed in G2019S and R1441G L2PD was done in Metascape under a multiple testing adj. P<0.05, here denoted as a dashed red line. (**a**) GO enrichment plot in G2019S L2PD vs controls. (**b**) GO enrichment plot in R1441G L2PD vs controls. Proteome changes related to both mutations showed affection of similar functional terms affecting the endolysosomal pathway (red asterisks), protein homeostasis (green), and mitochondria function (blue).

**Suppl. Fig. 4.**
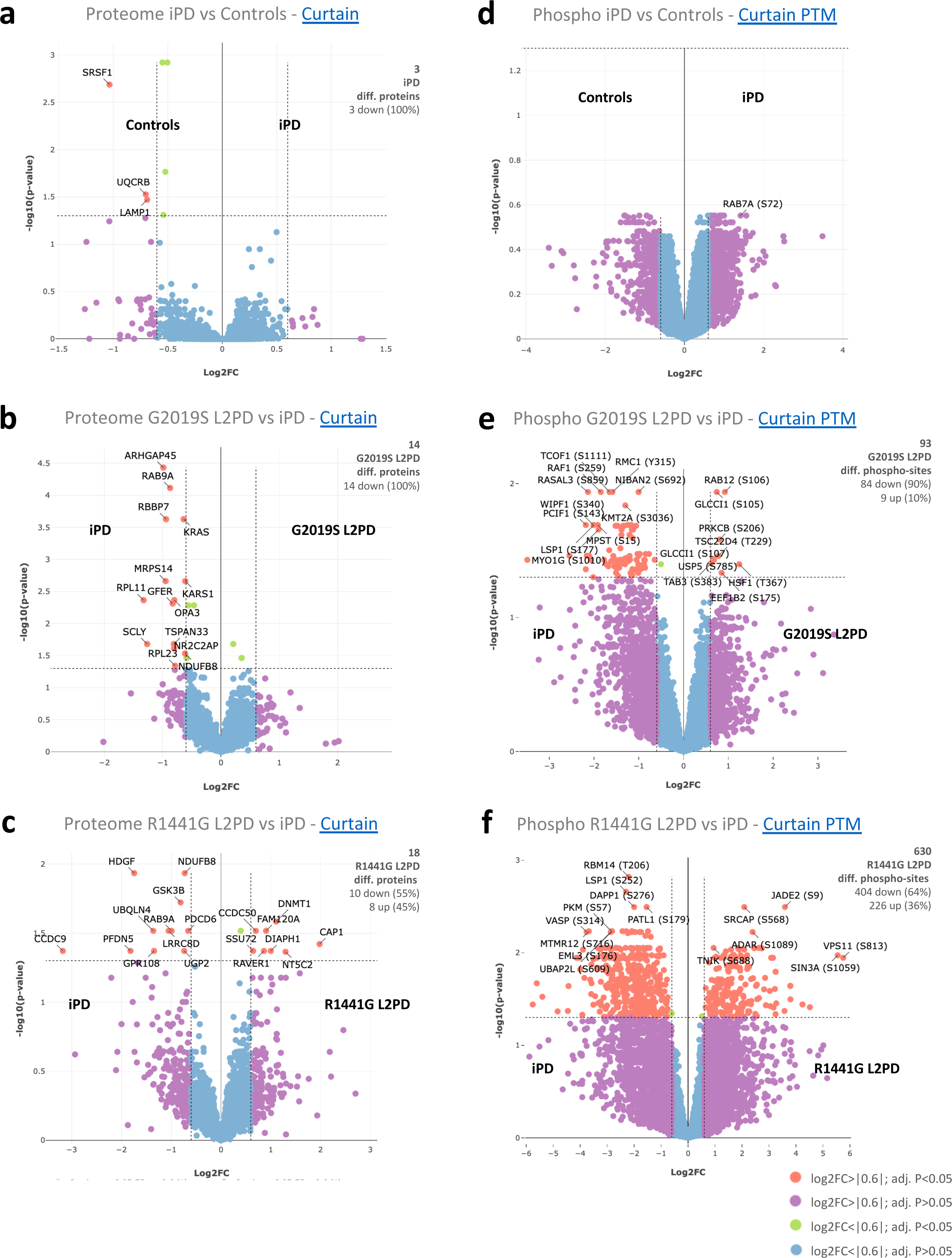
Proteome and phospho-proteome analysis of iPD compared to L2PD. Curtain weblinks provide access to raw and differential analysis data. The legend colour code shows hits categorisation based on statistical significance and applies to all the panels. (**a**) Volcano plot of the proteome analysis in iPD vs controls showing no differential hit under the statistical cut-off used (<u>Curtain</u>). (**b**) Volcano plot representing protein differences between iPD and G2019S L2PD, showing iPD up-regulated proteins as red dots on the left (<u>Curtain</u>). (**c**) Volcano plot representing protein changes between iPD and R1441G L2PD, with iPD up-regulated proteins as red dots on the left, and iPD down-regulated (i.e., up-regulated in R1441G L2PD) as red dots on the right (**d**) Volcano plot of the phospho-proteome analysis in iPD vs controls showing no differential hit, despite being iPD the groups with larger sample size in the study (<u>Curtain PTM</u>). (**e**) Volcano plot representing phospho-protein differences between iPD and G2019S L2PD, with proteins hyper-phosphorylated in G2019S L2PD as red dots on the right, showing pSer106 RAB12 as top hit, and proteins hyper-phosphorylated in iPD (i.e., hypo-phosphorylated in G2019S L2PD) as red dots on the left (<u>Curtain PTM</u>). (**f**) Similar analysis as in the previous panel, here comparing the phospho-proteome comparison between iPD and R1441G L2PD (<u>Curtain PTM</u>).

**Suppl. Fig. 5.**
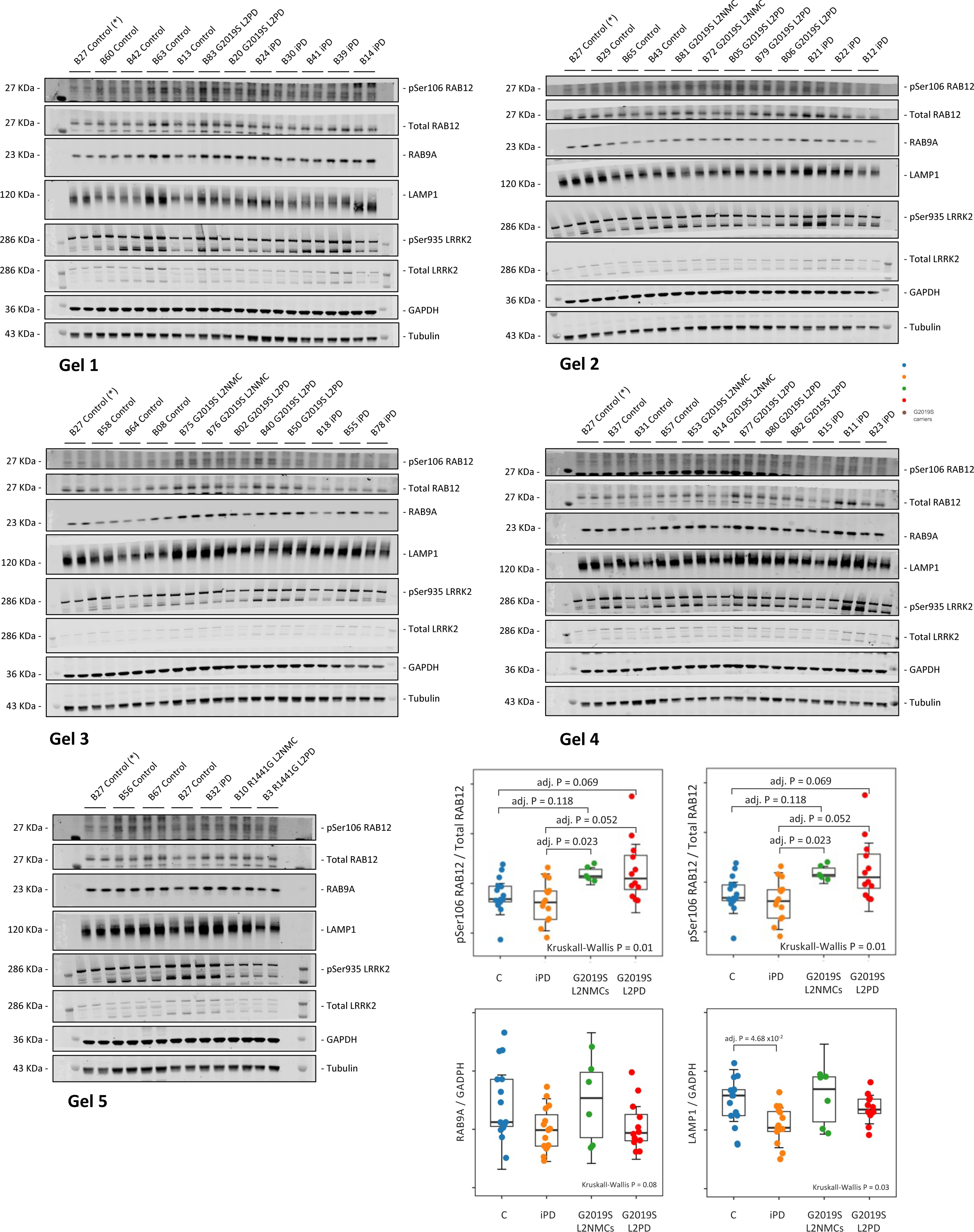
Expanded 1-year follow-up of pSer106 RAB12 and other markers by immunoblot. Full immunoblot assessment of pSer106 RAB12 phosphorylation levels, and expression levels of RAB9A and LAMP1, using >1-year follow-up PBMC samples from a subset of the LRRK2 cohort from Clínic-Barcelona (n=48), including G2019S L2PD (n=12), G2019S L2NMCs (n=6), iPD (n=15), and controls (n=15). Dot plots representing normalised levels after the band densitometric analysis for the various studied makers in all subjects studied in duplicates as it follows, pSer106 RAB12 / Total RAB12; RAB9A / GADPH; and LAMP1 / GADPH, all of them double normalised to the same intergel control also measured in duplicates. In each plot, overall intergroup differences were assessed using Kruskal-Wallis test followed by post-hoc Dunn’s test under an FDR multiple testing adjusted P<0.05. (*) Denotes intergel control.

**Suppl. Fig. 6.**
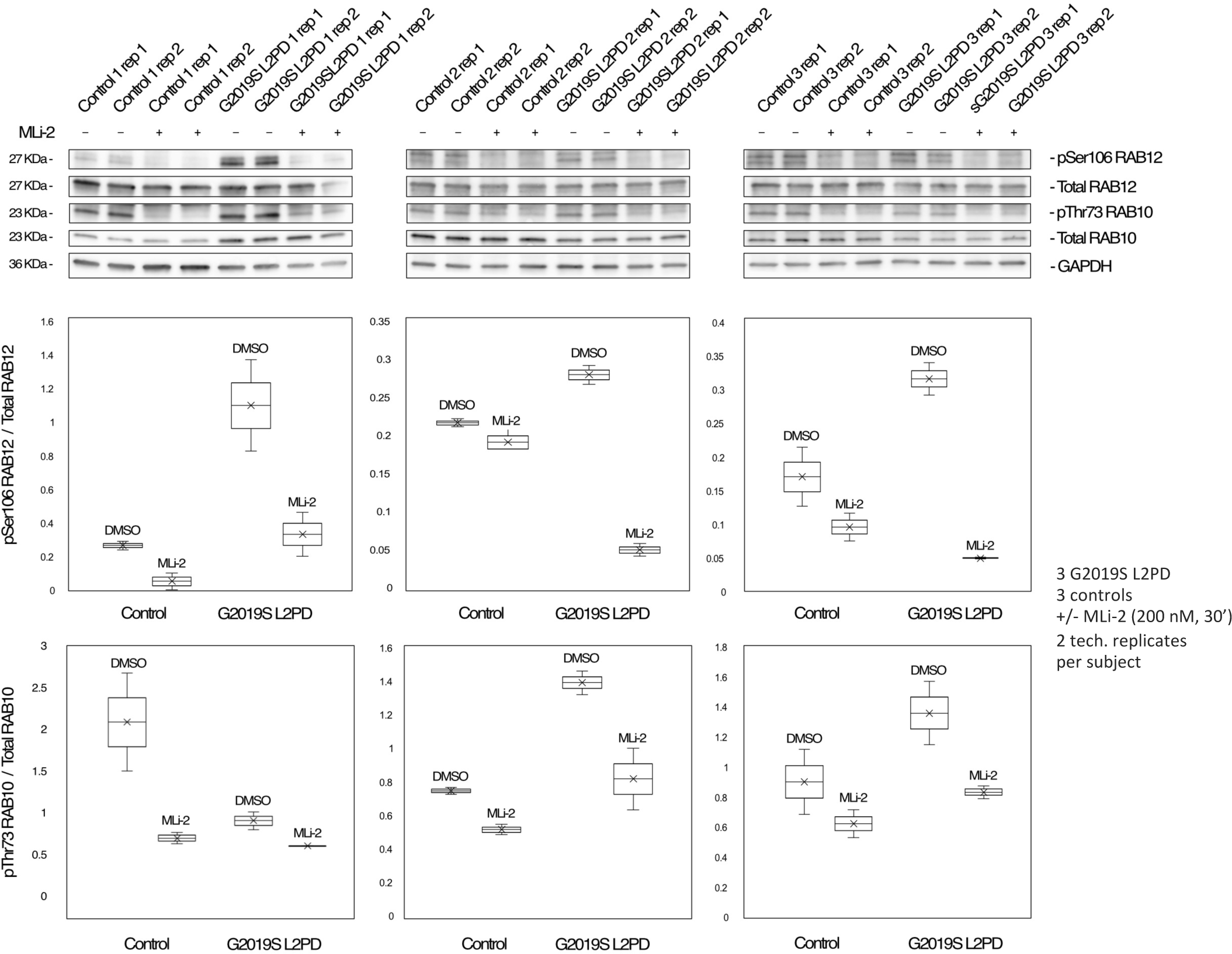
Expanded pSer106 RAB12 responsiveness to MLi-2 LRRK2 inhibition. Full immunoblot analysis of pSer106 RAB12 / Total RAB12 and pThr73 RAB10 / Total RAB10 using two technical replicates of PBMC lysates from G2019S L2PD (n=3) and healthy controls (n=3), treated with DMSO or the MLi-2 LRRK2 inhibitor (200 nM, 30 min), showing a diminishment of pSer106 RAB12 phosphorylation levels after LRRK2 inhibition by MLi-2 treatment.

**Suppl. Fig. 7.**
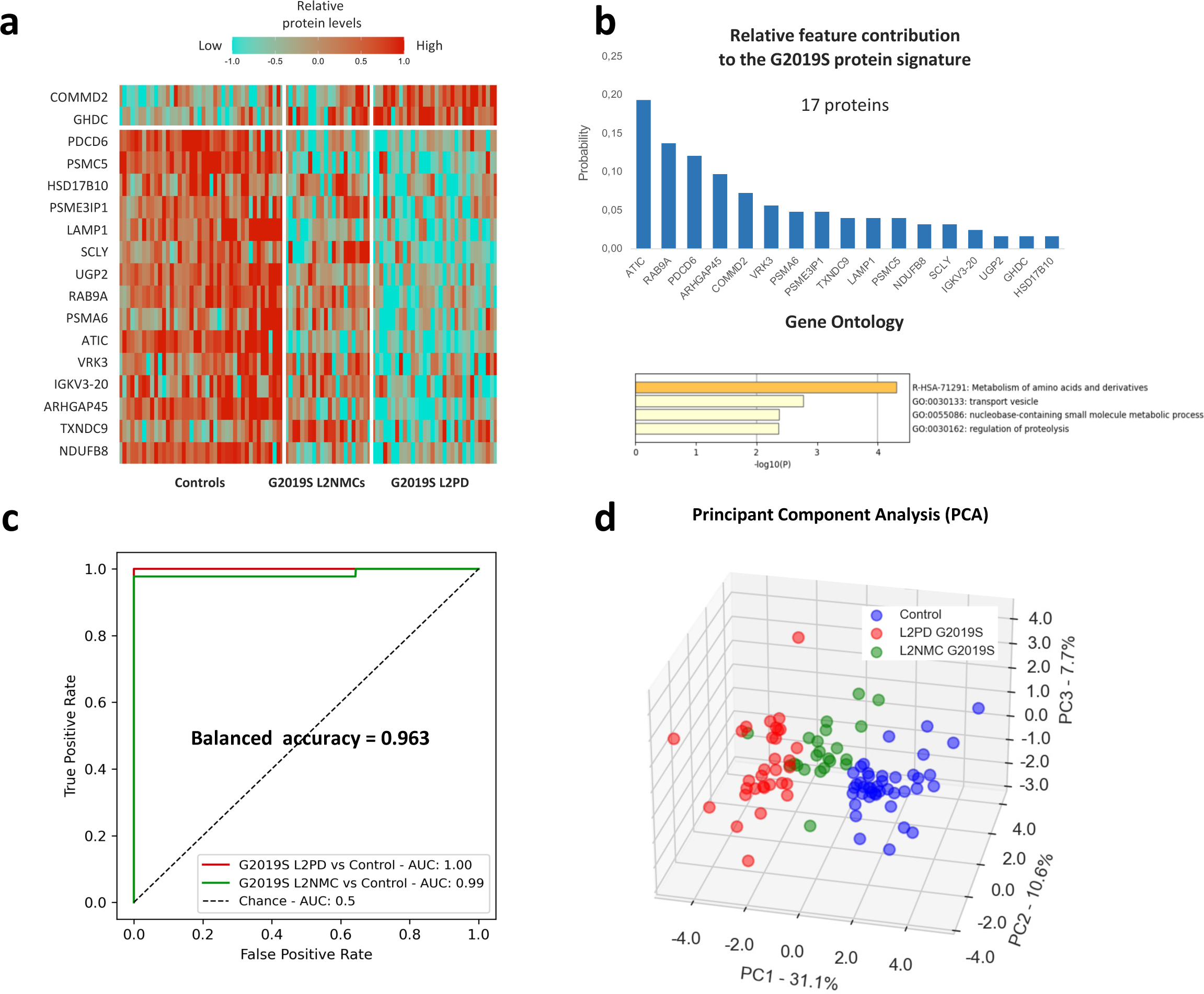
A 17-feature protein classifier for G2019S carriers. To explore alternative classifiers for G2019S carriers and healthy controls, here we applied the same method as described for the 18-feature G2019S phospho-/protein signature, and considered exclusively cross-group differential proteins but not phospho-sites. (**a**) 17-protein G2019S best classifier found in G2019S carriers, symptomatic and asymptomatic, and controls. (**b**) Relative contribution of features from the 17-protein G2019S classifier shown on the upper bar plot; Metascape gene ontology enrichment analysis of the 17-feature G2019S signature displayed on lower bar plot. (**c**) Receiver Operating Curve (ROC) analysis of the 17-protein G2019S signature with a balanced accuracy of 0.963 for discriminating G2019S L2PD, G2019S L2NMCs and controls, with an area under the curve (AUC) of 1.00 between G2019S L2PD and controls, and 0.99 between G2019S L2NMCs and controls. (**d**) Principal component analysis (PCA) based on the 17-protein G2019S classifier displaying different subject profiles of G2019S carriers and healthy controls based on LRRK2 mutation and disease status.

## ABBREVIATIONS

(Minimal abbreviations to be used in the text)

PBMCs: Peripheral blood mononuclear cells
CNS: Central nervous system
DIA: Data independent acquisition
MS: Mass-spectrometry
PD: Parkinson’s disease
LRRK2: Leucine-rich repeat kinase 2
L2PD: LRRK2-associated PD patients
L2NMCs: Non-manifesting LRRK2 mutation carriers
iPD: Idiopathic PD patients
p: Phospho-
FC: Fold-change

## Notes

### Competing Interest Statement

The authors have declared no competing interest.

### Funding Statement

This study was supported by the Michael J. Fox Foundation for Parkinson Research (MJFF) (MJFF 000858) to RFS, ME, CM, ES, JI, and JRM.

### Author Declarations

Probands participated in the study after local ethics approval and signed informed consent. The overall study was approved by the Ethical committee of the Hospital Clinic of Barcelona (code HCB/2020/1238).

